# PTSD therapy with fMRI-decoded neurofeedback bypassing conscious exposure: a randomized, double-blind, placebo-controlled study

**DOI:** 10.1101/2025.07.07.25330779

**Authors:** Toshinori Chiba, Kentaro Ide, Jessica E. Taylor, Aurelio Cortese, Takatomi Kubo, Keiichiro Nishida, Shuken Boku, Hiroyuki Toda, Miyako Shirakawa, Kaoru Amano, Hakwan Lau, Mitsuo Kawato, Ben Seymour, Ai Koizumi, Tetsufumi Kanazawa

## Abstract

Exposure-based therapy is effective for alleviating fear among patients with post-traumatic stress disorder (PTSD). Nonetheless, because the therapy itself induces fear, patients sometimes abandon treatment prematurely. One emerging alternative therapy employs real-time fMRI-Decoded Neurofeedback (DecNef). It aims to alleviate excessive physiological responses to threat, while bypassing conscious exposure procedure. With DecNef, neural activation patterns for feared cues are first identified. Then these patterns are subsequently induced through feedback without the patient’s awareness of the cues. However, evidence for DecNef’s efficacy is so far limited to laboratory settings and small numbers of patients. In the proposed study, we will test the effectiveness of DecNef for PTSD patients in a double-blind, randomized, placebo-controlled study with an improved procedure to decode nonconscious neural activities in response to trauma-related cues. To minimize patient variability, we will employ a crossover design with a six-month interval, considering the compatibility of DecNef to such a design. We will further examine the supposed implicit and stress-free nature of DecNef treatment. The mechanisms of DecNef will be examined with neuroimaging and computational approaches. If successful, this study may offer a less subjectively unpleasant new avenue for PTSD therapy.

## Introduction

Post-traumatic stress disorder (PTSD) is a debilitating disease that occurs following life-threatening traumatic events, and affects around 6.1 to 9.2% of the population sometime in life^1–4^. In typical clinical therapeutic approaches for PTSD, patients are repeatedly exposed to trauma-related stimuli. Accordingly, exposure-based therapy is often distressing for PTSD patients. In some cases, this causes them to re-experience the traumatic memories anew^5^, which can exacerbate their symptoms^6^. The reported dropout rate of exposure-based therapy is sometimes as high as 20-30% within a 2-month treatment period^7,8^, with some studies reporting even higher rates^9^. This rate is relatively higher than that including other psychological therapies for PTSD such as Present Centred Therapy, which have a dropout rate of around 16% (95% Confidence Intervals [CI] = 14-18%)^10^.

One recently proposed strategy to reduce PTSD symptoms without distress is to employ Decoded Neurofeedback (DecNef). DecNef estimates the likelihood that a target neural representation is present in a predefined brain region at a particular moment, through real-time decoding of functional magnetic resonance imaging (fMRI) signals^11–15^. By immediately presenting monetary reward feedback to the participants when the target neural representation is detected, this procedure reinforces participants to spontaneously induce the target neural activation patterns^16^. Importantly, participants typically remain unaware of the subjective content of the induced neural activation patterns^17–19^. As a result they do not consciously experience the relevant content and show no elevated physiological responses during the procedure^17,20^.

Previously, we leveraged the nonconscious nature of DecNef to reduce threat responses toward conditioned stimuli or feared animals at both physiological and neural levels^17,20,21^. In those studies, DecNef was used to reinforce participants to unconsciously induce the neural representations for feared stimuli with monetary reward. However, the evidence so far has been limited to healthy participants^20^, subclinical participants^17^, participants with specific phobias^21^, and a small number (N = 4) of PTSD patients (in a part of our pilot study^22^). Here, we propose to directly investigate the applicability of DecNef to alleviate PTSD clinical symptoms.

Specifically, we will test two primary and three secondary hypotheses related to the therapeutic effects of DecNef (Hypotheses **T**hera**p**eutics **e**ffect, **H_TPE_**). As primary hypotheses, we will test whether DecNef yields greater pre-to-post symptom reduction than control DecNef which induces fear-irrelevant stimuli (**H_TPE_ 1**). Second, we will assess the magnitude of this reduction, and test if DecNef can lead to a change in symptoms that exceeds a clinically meaningful threshold, defined as 10-point reduction on the clinician-administered PTSD scale (CAPS)^8,23^ (**H_TPE_ 2**). Additionally, our secondary hypotheses are that: DecNef’s therapeutic effects on PTSD symptoms persist for 2 months relative to the control DecNef (**H_TPE_ 3a**), and the magnitude of this long-term effect passes the clinically meaningful threshold (**H_TPE_ 3b**). We also hypothesize that DecNef effects will lead to alteration of physiological threat responses (including skin conductance response, SCR, and amygdala responses: **H_TPE_ 4a**), which may covary with symptom reductions **(H_TPE_ 4b)**. Finally, we hypothesize that these effects will be specific to PTSD symptoms rather than to symptoms related to comorbid conditions (**H_TPE_ 5**).

Besides the therapeutic effects, three challenges need to be met to develop DecNef procedures for PTSD patients. First, induction of trauma-related neural representations needs to remain nonconscious and should not induce excessive distress for PTSD patients, in order to minimize dropout^10^. Although healthy participants reported little distress during DecNef^17,20^, trauma-related neural representations may more readily evoke unpleasant subjective experiences in patients^24^. We will directly test the following four secondary hypotheses on **D**istress during **N**euro**F**eedback (**H_DNF_**): First, we hypothesize that participants will remain unaware of the DecNef-induced trauma-related content (**H_DNF_ 1**). That is, when asked subsequently what is their strategy for earning rewards during the DecNef procedure, fewer than i) 10%—a threshold for very frequent side effects, and ii) 50%— half—of them will report using trauma-related imagery. Also, fewer than i) 10% and ii) 50% of them will correctly guess that the relevant brain representation that leads to reward during the DecNef procedure is in fact trauma-related. Further, we hypothesize that participants will not be able to accurately guess the order of the two neurofeedback conditions (experimental DecNef vs control) in a two-alternative-forced-choice question, resulting at chance level accuracy (50%) in the whole patient group (**H_DNF_ 2**). We also hypothesize that DecNef training will not lead to subjective distress levels (SUDS) higher than both a tolerable threshold level at 50 points^25^, and pre-training baseline levels (**H_DNF_ 3**). Finally, we hypothesize that dropout rates will be lower than 14-18%, which is the 95% CI in the pooled dropout rate in the meta-analysis of randomized controlled trial (RCTs) of PTSD psychotherapy in general^10^ (**H_DNF_ 4**). Evaluations of those hypotheses will examine DecNef’s feasibility as low-distress PTSD treatment.

The second challenge is to minimize conscious exposure to trauma-related stimuli, not only during DecNef training, but throughout the entire DecNef procedure including its preparatory decoder construction sessions. In the standard DecNef procedure^17,20^, a participant is repeatedly exposed to visual stimuli to obtain neural activity patterns to build a decoder prior to the DecNef training. The decoder then allows the real-time detection of the neural activation patterns representing the target visual stimuli during DecNef. The explicit decoder construction procedure was not problematic with healthy participants who were fear-conditioned to the visual stimuli only after constructing a decoder. However, this approach is less favorable for PTSD patients, because they are already fearful of the trauma-related stimuli before the decoder construction.

Two potential approaches can bypass conscious presentation of feared stimuli during decoder construction: hyperalignment and visual pattern masking. Using hyperalignment, Taschereau-Dumouchel et al. inferred fear-relevant representations for participants with subclinical levels of phobic symptoms from the neural activity patterns of “surrogate” participants^17,26,27^. By having both sets of participants view a large set of stimuli (3,600 images of 40 different animals and inanimate objects), a decoder for feared stimuli can be aligned from the neural activation patterns of surrogate participants to the participants with phobia^17^. Hyperalignment can thus build a decoder of feared stimuli for participants with phobia, even if participants with phobia do not directly view the feared stimuli. This approach may be appealing for certain clinical conditions such as specific phobias of animals^17^ where a target representation is readily defined by other fear-irrelevant representations unlike PTSD.

In our pilot study conducted on patients with PTSD^22^, we instead used continuous flash suppression (CFS), which is a form of a binocular visual pattern masking, to render trauma-related images less visible^28^ during decoder construction. Specifically, a trauma-related image was presented to participants’ non-dominant eyes while salient flashing Mondrian patches were presented to their dominant eyes. Our previous study achieved clinically significant improvement in PTSD symptom by using CFS to build a decoder from the neural activation patterns in the superior temporal sulcus (STS) for trauma-related face stimuli^22^. Considering this, we will employ CFS and directly test our hypotheses on distress during decoder construction (Hypotheses on **D**istress during **D**ecoder **C**onstruction, **H_DDC_**). These hypotheses are that: Participants will remain unaware of trauma-related content during the decoder construction with CFS (detailed in Supplementary), and SUDS ratings for decoder construction sessions will remain below both a tolerable threshold level at 50^25^ and their baseline level from before the decoder construction procedure (**H_DDC_ 1**). Together with the assessments of distress during DecNef training (Hypotheses **H_DNF_**), evaluations of Hypotheses **H_DDC_** will determine whether unconscious and low-distress features are maintained throughout the DecNef protocol, ensuring clinical feasibility.

Finally, the third challenge is to resolve the two candidate mechanisms underlying the fear alleviation effects of DecNef. One candidate mechanism is that DecNef recruits a similar neural circuits as exposure-based therapies by repetitively inducing neural representations of feared stimuli, where the ventromedial prefrontal cortex (vmPFC) suppresses the amygdala to extinguish fear^29,30^. However, our previous study showed that the fear reduction effect was larger among individuals who had less vmPFC involvement during DecNef training^20^. With a neuroimaging approach, this study suggested an alternative mechanism involving the reward circuits, potentially supporting counter-conditioning of feared stimuli with rewarding feedback^31^. To dissociate the two candidate mechanisms, a preliminary investigation^22^ adapted a computational approach, applying variants of Rescorla-Wagner models^32^ to the data obtained from non-clinical populations in previous DecNef studies^17,20^. While the exposure-based model outperformed the counter-conditioning model, the results remained inconclusive^22^ and may differ among PTSD patients^33^. In this proposed study, we will test our hypothesis on **N**eural **M**echanisms (Hypotheses **H_NM_**) by applying the same neuroimaging^20^ and computational^22^ approaches to the data obtained from PTSD patients. Specifically, we will conduct two neuroimaging analyses to test whether DecNef relies on unique neural mechanisms that rely on the reward circuits (caudate and ventral striatum) rather than the fear extinction circuits (vmPFC). First, multivoxel decoding analysis will be used to evaluate transmission of trauma-related neural representations from STS, the target region for DecNef neural induction, to reward versus fear circuits during DecNef (**H_NM_ 1a**). Second, a functional connectivity analysis will evaluate the connectivity of STS with the reward and fear circuits (**H_NM_ 1b**). In Hypotheses **H_NM_ 1a and H_NM_ 1b**, we predict that the engagement of reward circuits during DecNef, compared with fear circuits, will correlate stronger with post-DecNef symptom alleviation, supporting the mechanism of counter-conditioning over exposure/extinction. With computational approaches, we will further examine whether the model based on the counter-conditioning computations will better explain the PTSD symptom reductions compared to the model based on the exposure/extinction-based computations (**H_NM_ 2**). Such results would provide evidence for reward-circuit-based therapeutic effects of DecNef, suggesting a mechanism distinct from traditional fear-circuit-based therapies.

We will confine participants to female patients who have developed PTSD from male violence. Females generally have twice as great a risk of developing PTSD as males^34^, and victims of intentional trauma such as male violence show worse prognosis than those of non-intentional trauma, such as natural disasters^35–37^. Enrolment of female survivors of male violence would enhance homogeneity among participants to facilitate investigations of DecNef effects and mechanisms, as PTSD patients are otherwise typically heterogeneous due to diversity of trauma types^35,37^ and sex differences^35,38^. In addition, female victims of male violence share a common small set of stimuli associated with traumatic experiences, especially angry male faces. As DecNef typically induces neural representations of a single target stimulus, this patient group with a specific trauma-associated stimulus may especially benefit from DecNef training.

To further minimize across participants variability, we will use a crossover design where participants will undergo experimental and control DecNef sessions with a 6-month interval between. A previous crossover DecNef experiment has successfully up- and down-regulated subjective confidence within single subjects across two DecNef sessions delivered with only a one week interval between^39^, which indicates that crossover design is appropriate for DecNef procedures. Our successful pilot PTSD-DecNef study^22^ also partially employed the crossover design (see Supplementary Table 1). Considering that effects of neurofeedback could last for 2-5 months^13,40,41^, we will minimize the potential anterograde learning interference effects from the first session to the second session by extending the interval to 6 months. As PTSD symptoms typically persist longer than 6 months^8^, we expect that effects of the second DecNef session can be assessed without a floor effect even after such a long interval.

Lastly, we adopted a co-design approach where participants with PTSD from our pilot study contributed to development and refinement of the study design (https://www.medsci.ox.ac.uk/research/patient-and-public-involvement). This pilot data yielded encouraging results, specifically that (a) the proposed decoder could be constructed with minimal distress to participants with PTSD, and (b) PTSD symptoms could be reduced via our DecNef technique. Leveraging the co-design approach, we confirmed that the scanner environment, rather than the presented stimuli, caused distress to participants. To address this, we decided to allow participants to bring a trusted companion and compensate their travel expenses and efforts. In this pre-registered study, the participants from the pilot and another 58 participants will contribute their insight and experience of the study to help us map a subsequent pathway to feasible clinical applications.

Overall, in this proposed study, we will validate the Therapeutic effects (**H_TPE_ 1-5**) of DecNef on PTSD in a randomized, double-blind, placebo-controlled, crossover design with a 6-month interval period with 58 participants with PTSD. We will evaluate low-distress characteristics of DecNef during neurofeedback (**H_DNF_ 1-4**) and its preparatory decoder construction sessions (**H_DDC_ 1**). We will further explore the Neural Mechanisms (**H_NM_ 1, 2**) regarding how DecNef reduces PTSD symptoms in neuroimaging and computational approaches.

## Methods

### Ethics information

This study will be conducted with approval from the Institutional Review Boards of Osaka Medical and Pharmaceutical University and Advanced Telecommunication Research Institute International. Participants will provide written informed consent prior to each session. This study has been registered with the University Hospital Medical Information Network (UMINID: UMIN000028148).

### Co-design and pilot data

A co-design approach was adopted (https://www.medsci.ox.ac.uk/research/patient-and-public-involvement) in which patient partners with PTSD played a key role in developing the paradigm, in terms of experimental logistics, study environment, management of adverse events, design of the task interface, optimisation of patient comfort, and task instructions. The resulting paradigm presented here reflects several iterations of this design cycle, with patient experience (as part of an explicit patient co-led risk management strategy) a fundamental part of this process. These patient partners and new patients to be enrolled in the pre-registered study will be actively involved as an oversight panel through the duration of the study, and will jointly consider the outcome of the study when complete.

We conducted a pilot study on a total of seven female participants with PTSD (mean age 39.7, SD = 10.1), three of which participated in both experimental and control conditions. This pilot study included data from four participants with PTSD that were reported elsewhere^22^ and the procedure used for these four participants was the same as that used for the other (previously unreported) three participants. In brief, neural activation patterns for trauma-related male angry faces and trauma-unrelated happy female faces (see *decoding session* for detail) were induced via DecNef in the experimental and control conditions, respectively. Procedures of the pilot study were mostly identical to those of this pre-registered study with two exceptions. First, while the pilot study was conducted in a single-blinded manner, we will conduct the pre-registered study in a double-blinded, completely randomized manner. That is, experimental and control conditions in the pilot study were blinded only to participants whereas the pre-registered study will be further blinded to experimenters and analysts. The pre-registered study will thus validate that the observed effect in the pilot study is not due to a placebo or experimenter effect. Second, we mainly used mrVISTA and TurboBrainVoyger instead of Statistical Parametric Mapping (SPM; Welcome Department of Imaging Neuroscience, London, UK) to analyze the pilot fMRI data, consistent with our previous studies^20^ (see Supplementary Information for details). In the proposed study, we will use more commonly used software, SPM12, for ease of DecNef applications in clinical settings (see Design section for details).

In our pilot study, we observed that participants in experimental conditions (N = 6) showed improvement in symptom severity relative to participants in control conditions (N = 4) (Figure 1). Symptom severity was measured using the clinician-administered PTSD scale for DSM-IV, 1-wk Symptom Status Version (CAPS-SX^25^), which is the gold standard in this field. On average, symptom severity assessed by CAPS^25^ was reduced by 35.8 points (SD = 8.8) in the experimental group from the pre-training test (−1 day) to the post-training test (+1 week). Meanwhile, only 4 points (SD = 21.1) were reduced in the control condition group. As 10-point changes in CAPS are generally considered clinically meaningful^8,23^, we concluded that those experimental conditions are worth pursuing in the pre-registered study. The alleviating effects of DecNef were consistent across all symptom clusters of PTSD defined in the DSM-IV: Re-experiencing symptoms was reduced by 15.3-points (SD = 6.3) in the experimental group and by 3-points (SD = 6.4) in the control group. Avoidance symptoms were reduced by 12.7-points (SD = 7.6) in the experimental group and by 0.75-points (SD = 11.9) in the control group. Hypervigilance symptoms were reduced by 7.8-points (SD = 3.4) in the experimental group and by 0.25-points (SD = 6.7) in the control group. Using pilot data, a two-tailed t-test comparing total CAPS score reduction from the pre-training (−1 day) to post-training (+1 week) yielded significant results (t (8) = 3.4, *p* = 0.01, Cohen’s *d* = 1.97, 95% CI: 10.0-53.7).

**Figure 1.**
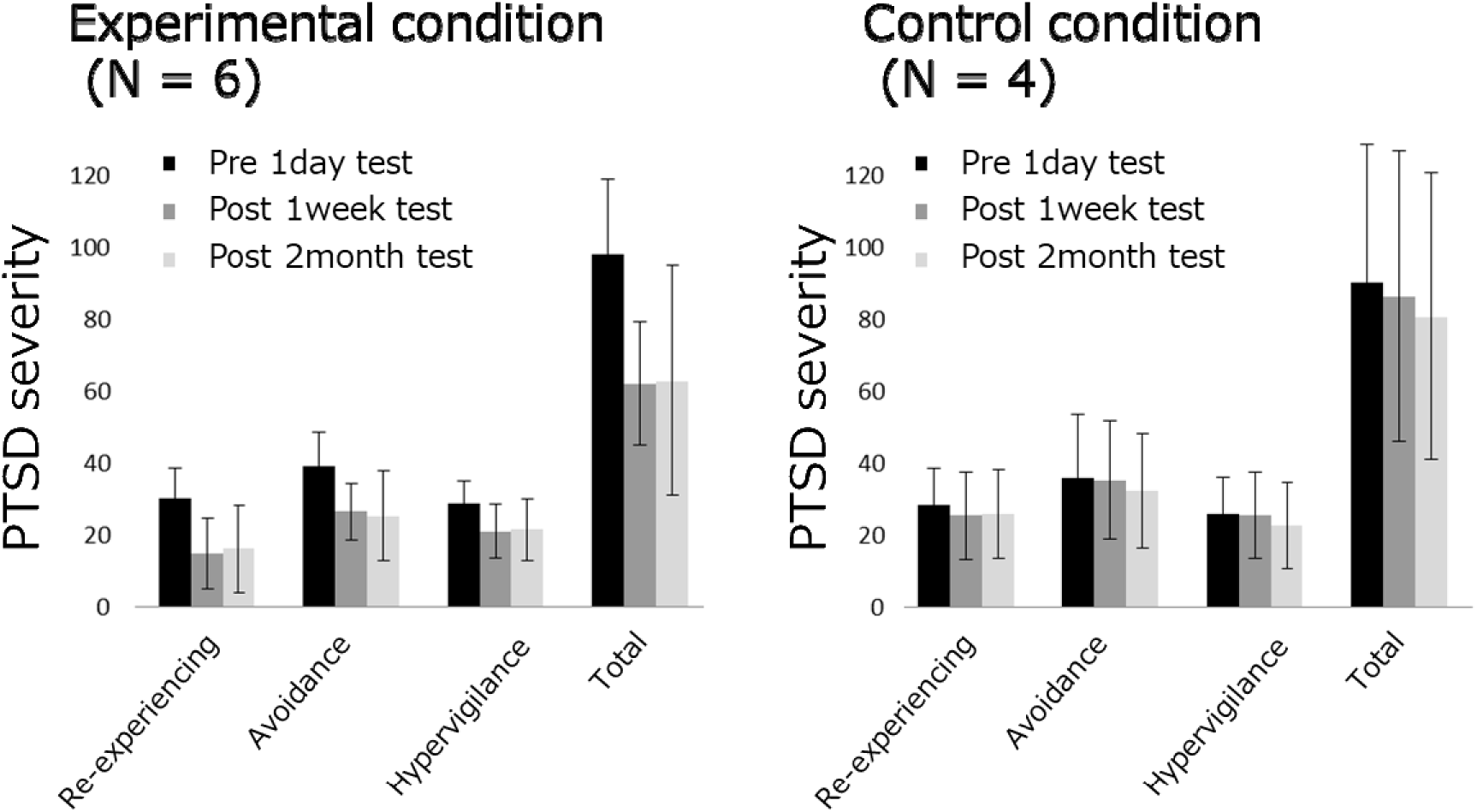
DecNef effects in the pilot study. Mean PTSD severity, as measured with CAPS scores, for three PTSD symptom clusters and their total in each test session. Error bars represent standard deviations.

In qualitative interviews after the entire pilot procedure, some participants reported a state of distress unrelated to DecNef itself, i.e., distress in traveling to the institute or distress in entering the confined space of an fMRI scanner. Further, one out of ten participants used a strategy related to her trauma during manipulation of brain activity in the first experimental session out of six sessions (three experimental sessions and three control sessions), during which she had higher distress. However, she used a different strategy unrelated to trauma in the remaining five sessions, which involved no distress, as revealed during the post-experiment qualitative interview. All participants, including this one participant, reported that the decoding and DecNef procedures were generally endurable. Further, all participants reported that they had no idea as to the content of target neural representations. Beyond laboratory assessments, we observed real-life improvements of patient symptoms. Because of their symptoms, two of six participants were unemployed at the time of enrollment in the experimental group. After participating in the experiment, however, these two were able to start new jobs for the first time in one year and ten years, respectively.

### Intended Endpoint

The primary endpoint of this study is PTSD severity reduction, measured by CAPS-SX^25^, as in our pilot study. Consistent with previous studies, 10-point changes in CAPS will be considered clinically meaningful^8,23^. The secondary endpoints are: 1) changes in physiological threat responses to threats (angry faces), as measured by SCRs and amygdala reactivity, and 2) reduction of PTSD, depression, and anxiety severity, as measured with validated self-administered questionnaires: the Impact of Event Scale-Revised (IES-R)^42^, BDI^43^, and STAI-Y^44^, respectively. These additional psychological assessments will be included to examine whether the effects of DecNef are specific to PTSD symptoms.

Objective threat responses will be assessed to determine their association with changes in CAPS scores that rely on subjective reports. Our objective measurements will include SCRs and amygdala reactivity, which are gold standards to index physiological threat responses. Alterations in the levels of both measures are pronounced during PTSD development^45,46^. However, while elevated physiological threat responses (including SCRs and amygdala reactivity) are usually associated with higher overall PTSD severity, lower responses are also known to accompany greater dissociative symptoms of PTSD^47–51^. Given that both elevated and reduced threat responses signify the severity of PTSD, we do not make any predictions regarding directionality of changes in physiological threat responses following DecNef. As an exploratory analysis, we will minutely examine associations between physiological measures, subjective PTSD symptoms, and PTSD subtype (see Supplementary Information for details).

### Design

#### Design overview

We will test the effectiveness of DecNef for patients with PTSD in a double-blind, randomized, placebo-controlled crossover study. First, all participants will undergo one preparatory decoding session where neural representations for angry faces and happy faces will be identified for each patient. Second, they will undergo session blocks for the *experimental* and *control* conditions of DecNef, in a counterbalanced order (Figure 2). Each session block consists of three consecutive days of DecNef sessions. In the experimental condition of the DecNef session, neural representations of trauma-related male angry faces will be induced as target activation patterns. In the control condition, neural representations of trauma-unrelated happy female faces will be the target of induction. In each session, target neural representations (angry male or happy female faces) will be reinforced with monetary rewards on three consecutive days (Figure 2). The effect of each condition will be evaluated in one pre-training test (−1 day from DecNef) and three post-training tests (+1 day, +1 week, and +2 months). We will allow a maximum of 2 days of jitter for post-training test (+1 week) and 2 weeks of jitter for post-training test (+2 months), depending upon participant schedules or requests, or MRI malfunctions. Decoding and the first DecNef session block will be separated by a few days to a few weeks. The second DecNef session block will be conducted more than 6 months after the first DecNef session block (i.e. more than 4 months after the final post-training test of the first DecNef session block).

**Figure 2.**
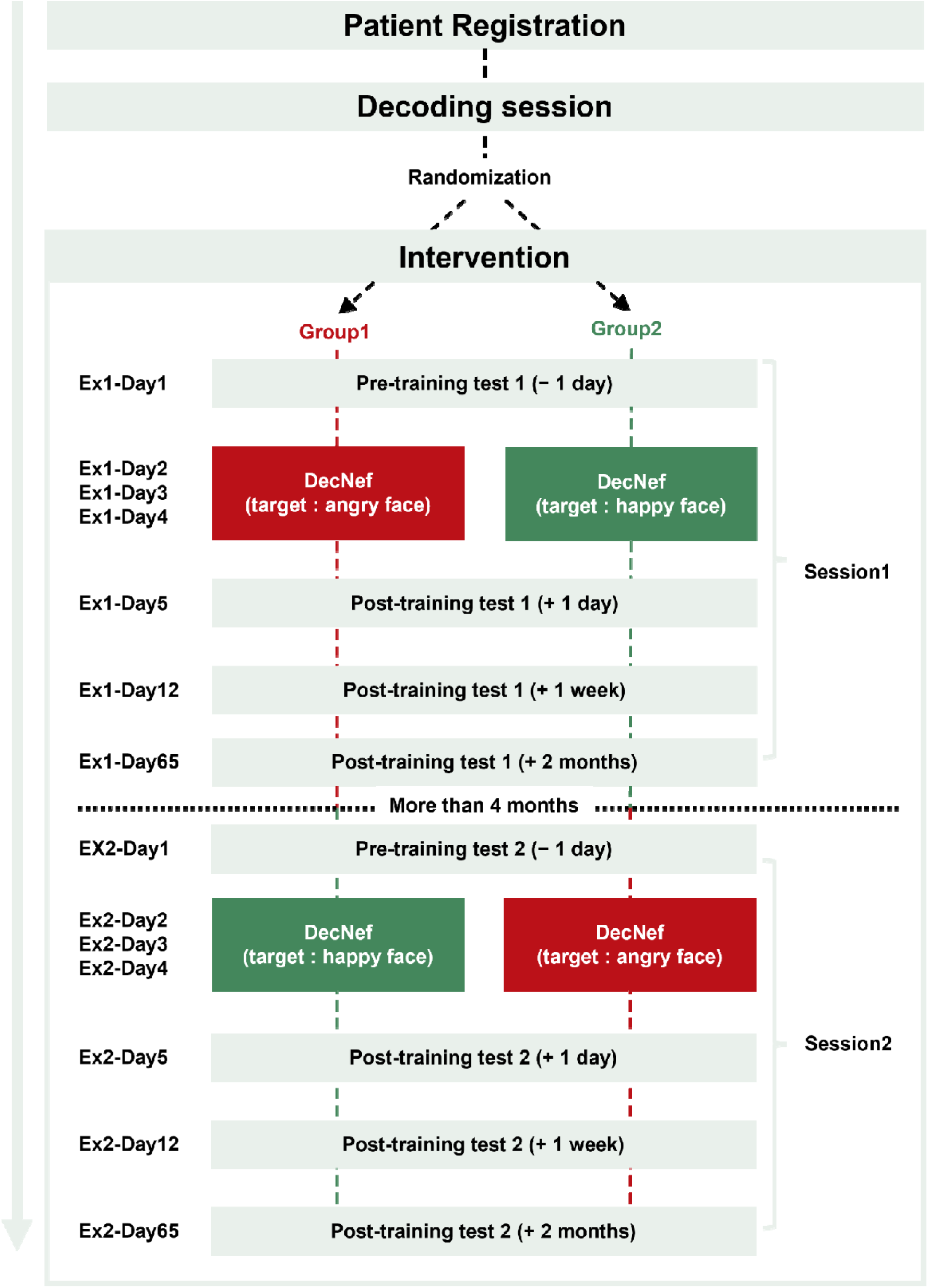
Overview of experimental design. Fifty-eight participants with PTSD will perform DecNef training in both the experimental and control conditions. The effects of DecNef in each condition will be assessed with pre- and post-training tests. The order of conditions will be double-blinded and semi-randomized for each patient.

#### A double-blind, randomized, placebo-controlled crossover design

In addition to a typical double-blind design where conditions are blinded to the participants and experimenters, evaluators and analysts will also remain blind. To achieve this, before starting the experiment, a correspondence table of IDs and conditions will be created by a program and kept in a state that only the program can reference. The experimenter will then assign IDs to the participants without accessing the assigned conditions. During DecNef sessions, the execution program will refer to the correspondence table to apply the conditions corresponding to the IDs. Likewise, the analyst will refer only to the IDs, and the analysis execution program will automatically reference the correspondence table internally to distinguish between the two conditions. The correspondence table will be maintained and verified only by a non-author monitoring personnel, and researchers will access the table only after completing all the analyses.

Half the participants will undergo the experimental condition first and another half will undergo the control condition first (see Supplementary for the details). A computer program will semi-randomly assign the condition order for a given participant. To ensure similar levels of PTSD severity between the condition order groups prior to the enrollment, the program will assign participants to approximately equate the proportion of moderate PTSD (45≤CAPS ≤ 60; M-PTSD) and severe PTSD (CAPS > 60) cases.

We will use trauma-unrelated neural representations—those for happy female faces—as a placebo in the control condition instead of sham or yoked feedback, which typically relies on signals regardless of the participants’ performance, for four reasons. First, it is less ethical to require patients to engage in seemingly ineffective procedures albeit their significant effort and time. Second, if both the experimental and control conditions reduce distinct PTSD symptom clusters as hypothesized (see “Analysis of DecNef effects on each PTSD symptoms cluster” in the Supplementary Information), we can evaluate distinct effects across conditions within participants using a crossover design. Third, we observed that the control condition is less effective than experimental conditions in our pilot^22^. Fourth, this approach prevents participants from guessing the assigned condition based on the controllability of brain activity.

The crossover design can cancel across participants variability since each participant will undergo both conditions sequentially. To minimize a potential carryover effect from the first to second condition, we will insert a 6-month interval between them. Carryover effects, if any, should last at most for 6 months as a previous ROI-based neurofeedback study reported symptom reduction that lasted several months maximum^40^. Similarly, effects of DecNef or functional connectivity based neurofeedback may also last several months^13,41^. While anterograde effects were reported in previous DecNef experiments^39^, Cortese et al. successfully induced bi-directional behavioral changes by reinforcing two opposing neural representations in two session blocks with just one week interval^39^. Thus, an interval of 6 months is likely to minimize the anterograde effect from the first training.

One may worry that most participants might no longer exhibit PTSD symptoms when they enter the second DecNef session block after the 6-month interval. However, this is unlikely since PTSD patients typically remain symptomatic (total CAPS score > 20^52^) even after several years of spontaneous observation or following post-trauma-focused interventions^53,54^. Women with PTSD, the target population in this proposed study, have been shown to remain symptomatic (CAPS > 45 at the lower boundary of the mean) 6 months after trauma-focused psychotherapy, as demonstrated in a large randomized controlled trial (N = 284)^8^.

#### MRI parameters and equipment

Experiment will be conducted using a 3.0□T scanner (Prisma; Siemens, Erlangen, Germany) with a 32-channel head coil at the ATR Brain Activation Imaging Center, using parameters identical to those used in Koizumi et al. and Taschereau-Dumouchel et al^17,20^. Specifically, we will scan 33 interleaved axial slices, 3.5-mm thick, without a gap, parallel to the anterior and posterior commissure line, with a T2*-weighted gradient, echo-planar imaging (EPI) sequence [repetition time (TR)□=□2000□ms, echo time (TE)□=□30□ms, flip angle (FA)□=□80°, field of view (FOV)□=□192□× 192□mm^2^, voxel size□=□3□×□3 ×□3.5 □mm^3^]. During the scan, we will record SCR from distal phalanges of the index and middle fingers of the left hand using BrainAmp Ag/AgCl sintered MR electrodes (Brain Products, inc.). For anatomical reference, we will acquire high-resolution T1-weighted images of the whole brain from all participants using a magnetization-prepared, rapid-acquisition gradient echo sequence (MPRAGE) [TR□=□2250□ms, TE□=□3.06□ms, FA□=□9°, FOV□=□256□×□256 mm^2^, thickness = 1 mm, 0 mm slice gap, voxel size□=□1□×□1□×□1□mm^3^].

#### Decoding session

The aim of the decoding session is to obtain fMRI data for constructing a multivariate pattern analysis (MVPA) decoder to classify activation patterns evoked by trauma-related angry male faces versus unrelated happy female faces. Based on previous studies reporting that the superior temporal sulcus (STS) contains information about the category of perceived emotions in faces^55,56^, the STS was selected as a region of interest (ROI) for our MVPA (see Decoder construction and Online fMRI analyses during DecNef). The decoder will be used in subsequent DecNef sessions to evaluate the trial-by-trial likelihood that current activation patterns in participants represent trauma-related angry male or unrelated happy female faces in experimental or control conditions, respectively. Here, we will refrain from using specific faces of actual trauma perpetrators for a given participant since such an image is not always available in future clinical settings. Instead, we will use grayscale pictures of four angry male faces and four happy female faces from the ATR Facial Expression Image database (DB99) where recognizability of intended emotions is well-validated (ATR-Promotions, Kyoto, Japan, http://www.atr-p.com/face-db.html).

A modified CFS method will be applied to render face presentations less conscious and distressful. Because decoding brain activity patterns for invisible visual stimuli are likely more difficult^57,58^, we will customize the CFS method to achieve an optimal balance between high decoding accuracy and conscious awareness of stimuli by gradually reducing mask contrast.

In each trial, a face stimulus (angry male or happy female) will be gradually faded into the non-dominant eye by linearly increasing its contrast over 1 s and then remaining constant for 6 s, while CFS masks containing salient Mondrian patches will be flashed to the other eye for 7 s at 10Hz. The mask will be presented at the same contrast as the face images for the first 1 s and then gradually faded-out linearly over 6 s. These presentations will be followed by presentation of a fixation disc to both eyes (for 7 s). We will extract preprocessed fMRI signals from the 6 s of each trial where a face is presented with a constant contrast. Because the TR = 2 s, this will provide three data points per trial, which will be averaged for each voxel from STS subregions and then used to construct a decoder to classify activation patterns for trauma-related angry male versus unrelated happy female faces. The session is subdivided into 11 runs of 17 trials (5 min) each. Sixteen exemplars (eight angry male faces and eight happy female faces) will be shown once per run in a randomized order. Each run starts with a dummy trial where a randomly selected happy female face is presented to capture irrelevant physiological threat responses due to the orienting effect^59^. This dummy trial will be discarded from subsequent analyses. Thus, the session comprises a total of 187 trials (88 trials per face category and 11 dummy).

To assess the level of conscious awareness of masked face images, participants will be told to press a button when they see any image other than Mondrian images during MRI scan. Trials where the button is pressed/unpressed will be defined as conscious/nonconscious trials, respectively. We will not impose further questions to confine the content of images reaching consciousness, as such questions themselves could alter prior expectations for presented images, which would in turn modulate actual neural activations^60^ to interfere with decoding. Before the beginning of the decoder session, eye dominance will be examined using the hole-in-a-card test^61^. T1-weighted images will be acquired in this session and used as anatomical references for functional images from all sessions.

To assess the level of distress during decoding, participants will be asked to rate the subjective units of discomfort (SUDS^25^) that they experienced during the decoding session, as well as before the first fMRI of the decoding session, as a baseline. The SUDS scale is a continuum from 0 (no distress) to 100 (maximum load), and 50 represents the strongest load that is still considered endurable. SUDS will be assessed at the end of each fMRI run, roughly every 5 min. This is comparable to a previous study that assessed SUDS during prolonged exposure therapy^62^. To avoid imposing task pressure on participants to report extra discomfort due to frequent assessments, we will add a dummy question on sleepiness level (Stanford sleepiness scale) following each SUDS assessment.

#### DecNef session

The aim of DecNef sessions is for participants to repetitively induce target STS activation patterns, specifically those for trauma-related angry male faces in the experimental condition and those for the trauma-unrelated happy female faces in the control condition. To achieve nonconscious induction of target activation patterns, we will neither give explicit instructions on how to manipulate their brain activity nor disclose the identity of the target patterns. We will reinforce participants with larger monetary rewards for inducing brain activity patterns more similar to the target patterns. In each session block, DecNef sessions will be conducted on three consecutive days. On each day, participants will engage in up to 12 fMRI runs (one run = 350□s), which are separated by brief break periods upon participant request. Each run consists of an initial 30 s fixation period followed by 16 trials (one trial = 20□s). To avoid unsaturated T1 effects, we will discard fMRI data for the initial 10 s of the fixation period. We will instruct participants to fixate their eyes on a dot presented at the center of the display throughout a run (Figure 3). Each trial involves the following sequence: an induction period (6 s), a fixation period (7 s), a feedback period (1 s), and an inter-trial interval (6 s) (Figure 3). During the induction period with a green dot presented centrally on-screen, participants will be instructed to maximize the size of the gray disc (surrounding the green dot) that serves as feedback by somehow manipulating their brain activity. Subsequent to the induction period, the fixation target will be presented as a central white dot on-screen, and the STS activation pattern during the induction period will be analyzed online to estimate the likelihood of target activation patterns (see Online fMRI analyses during DecNef). Then, feedback will be presented as a gray disc, with a radius proportional to the average likelihood (ranging from 0 to 100%) of target face representation in the STS during the induction period of the current trial. Participants, importantly, will not be informed of the association between the induced neural representations, e.g., an angry face, and the size of the disk itself.

**Figure 3.**
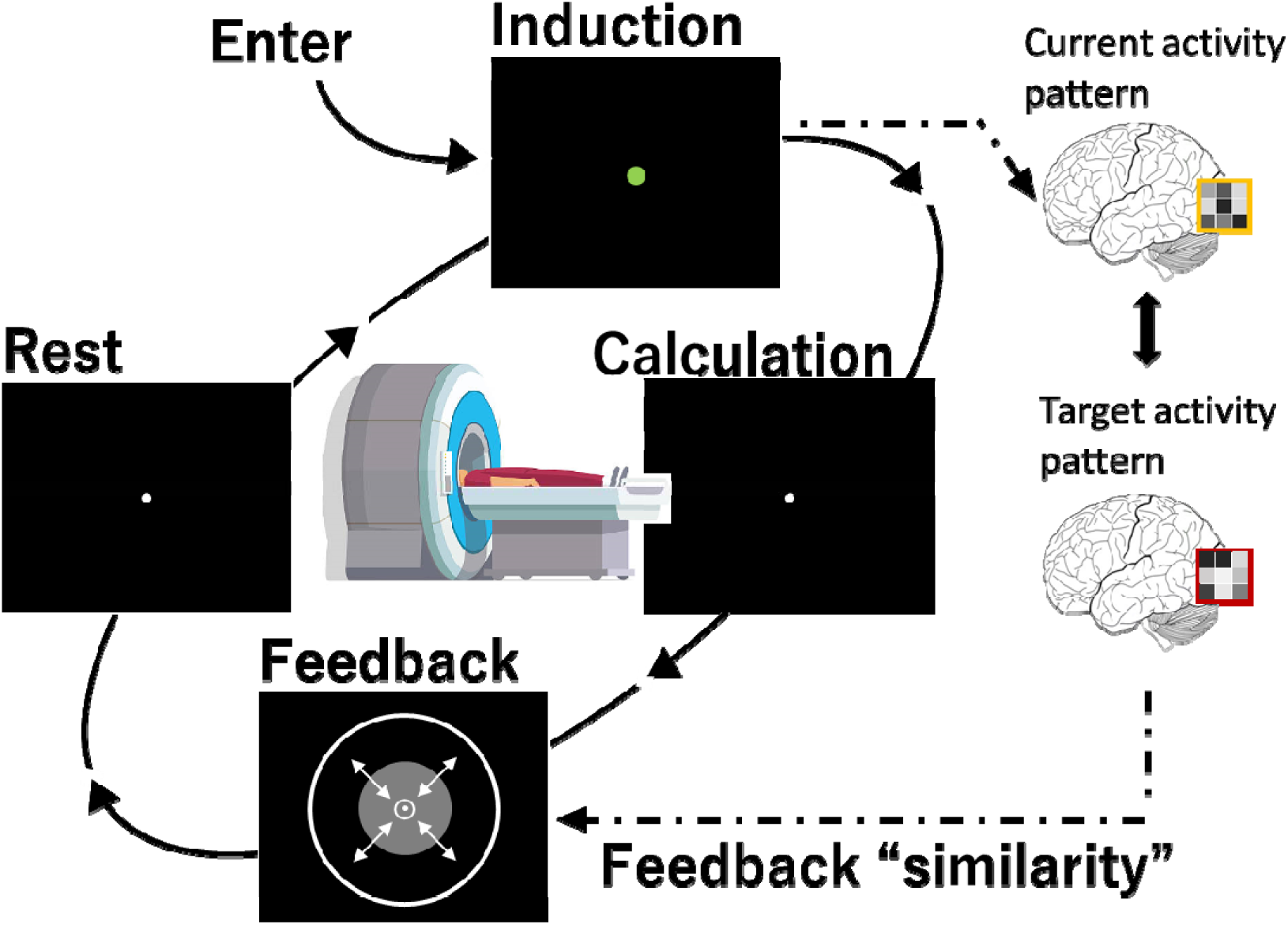
Overview of DecNef session. Each trial during DecNef sessions will begin with an induction phase, in which participants will manipulate their brain activity patterns to approach a predetermined target activity pattern. In the calculation phase, similarity between current brain activity patterns and the target activity patterns (angry or happy faces) will be calculated as likelihoods of the target representation using a pre-trained, multivoxel decoder. Calculated similarity will be fed back to the participant in the feedback phase. After the rest phase, participants will enter the induction phase for the next trial. By repeating trials, participants will be reinforced to induce the target brain activity patterns through trial and error.

In addition, at the end of each DecNef session block (Day 4 for the first DecNef session block; Day 184 for the second DecNef session block), participants will write the strategy they used during the induction period when they were manipulating their brain activity to maximize the size of the gray disc. Participants will then be asked to respond to a four-forced-choice question: “Do you think you were guided to induce in your brain a representation of 1) mechanical tools, 2) animals, 3) a smiling woman, or 4) an angry man.” In addition, a two-forced-choice question will be presented at the end of all procedures, i.e., the last test following the second session block, as follows: “You participated in two conditions in randomized order. Experimenters tried to induce representations in your brain of angry male faces and happy female faces. Which condition do you think you received first? (choose either angry male faces or happy female faces condition).” The participants will not be provided with any feedback, which minimizes the risk that these procedures interfere with the therapeutic effects.

To assess the level of distress during DecNef, the SUDS will be assessed at the end of each fMRI run, roughly every 5 min, as well as before the first fMRI run of each DecNef session as a baseline. A dummy question on sleepiness level (Stanford sleepiness scale) following the SUDS assessment will also be added, as in the decoding session. In addition, a two-forced-choice question will be delivered at the end of all procedures, i.e., the last test following the second session block, as follows: “Which DecNef session block was more discomforting: the first or second DecNef session block?”.

#### Pre/Post test sessions

We will assess participant PTSD severity, depression severity, anxiety severity, and the strength of their physiological threat responses at the following times for each DecNef session block: the day before the first day of DecNef (pre-training test (−1 day)), and one day after (post-training test (+1 day)), one week after (post-training test (+1 week)), and 2 months after (post-training test (+2 months)) the last day of DecNef (Figure 1). The procedure for all of these tests will be mostly identical. Each test will generally assess subjective PTSD severity as measured using CAPS, physiological threat responses to threat (angry faces) as measured by SCRs and amygdala reactivity, and subjective severity of other psychiatric symptoms as measured by questionnaire-based psychological assessments (see section *Endpoint*). One exception is that the CAPS will not be assessed during the post-training test (+1 day). This is because CAPS-1-week assesses PTSD severity over the past week, which means that at least one week should lapse after the pre-training test to avoid an overlap in periods. Therefore, changes in PTSD severity from before until after DecNef will be assessed using the CAPS at the pre-training test and the post-training test (+1 week). Further, long-term effects of DecNef will be assessed using the CAPS at the post-training test (+2 months).

In each test, physiological threat responses to angry male faces and happy female faces will be measured simultaneously via SCRs and amygdala reactivity inside an MRI scanner. The procedure of the MRI experiment for pre/post-training test sessions is consistent with that of the decoding session, except for the following two points. First, CFS masks that render the target face less visible will be presented at the same contrast as the target face without gradually decreasing their contrast, while the target face is presented to the other non-dominant eye. While the CFS masks may somewhat suppress SCRs and amygdala reactivity, we prioritize minimization of subjective distress during the test. Second, the whole experiment will comprise two runs, instead of 11 runs as in the decoding session.

### Sampling plan

#### Sample size

We will conduct this study at Advanced Telecommunications Research Institute International (ATR), Japan, with a total of 58 participants with PTSD between 20 and 55 years old. **H_TPE_ 1 (Primary Outcomes):** We compared two sample size estimates to select the larger sample size, avoiding potential underestimation due to sole dependence on a single pilot study or on previous literature^63,64^. In the first sample size estimation, we calculated the effect size using the SIMR^65^ in R based on the pilot data. The SIMR can estimate the sample size of the mixed-effects model planned in this study, based on the simulated or estimated coefficients and variability. Here, we used coefficients and variability estimated from our pilot study^22^ to test the efficacy of DecNef on PTSD symptoms (also see ***Table 1. Design Table***), which revealed that a sample size of 20 for each condition is sufficient to achieve 95% power. In the second estimation, we conducted a power analysis based on the latest meta-analyses on the PTSD psychotherapy (including prolonged exposure)^66^. Here, since the coefficients and variability in the mixed-effects model are unavailable and highly uncertain, we used G*Power 3.1.9.7^67^ to calculate the sample size to detect the minimally clinical important differences as smallest effect sizes with a t-test. Based on the meta-analyses on the psychotherapy that the effect size of Hedge’s g =1.248 (95% CI: 0.684-1.813) in comparison with treatment as usual (or routine care) or effect size of hedge’s g = 1.524 (95% CI: 1.235-1.814) in comparison with wait list^66^, we assume 0.684 (the lowest bound) as the smallest effect size of interest (SESOI). With a two□tailed t-test (α = .05, β = .95), this calculation estimated that we need 57 participants to detect a significant effect. We will therefore enroll 58 participants (minimum number for counterbalancing) to complete all sessions. Experiments will be continued until 58 participants complete all procedures. For the efficacy, safety, futility, and time efficiency we will conduct interim analyses based on the O’Brien-Fleming stopping boundaries^68^, with predefined significance thresholds: the first analysis at 10% of the data (i.e., 6 participants) with *p* < 0.0005, and the second analysis at 50% of the data (i.e., 29 participants) with *p* < 0.015. The trial will be terminated early if these thresholds are met for both of primary outcomes (i.e., **H_TPE_ 1 and 2**). Considering the long interval period of more than half a year, we will continue enrolling participants toward the full study target (i.e., N = 58) until the stopping rules are met. If the stopping rules are met, we will halt further enrollment while allowing already enrolled participants to complete the full study procedures. All analyses will be conducted using data from participants who have completed the full procedures.

**Table 1.**
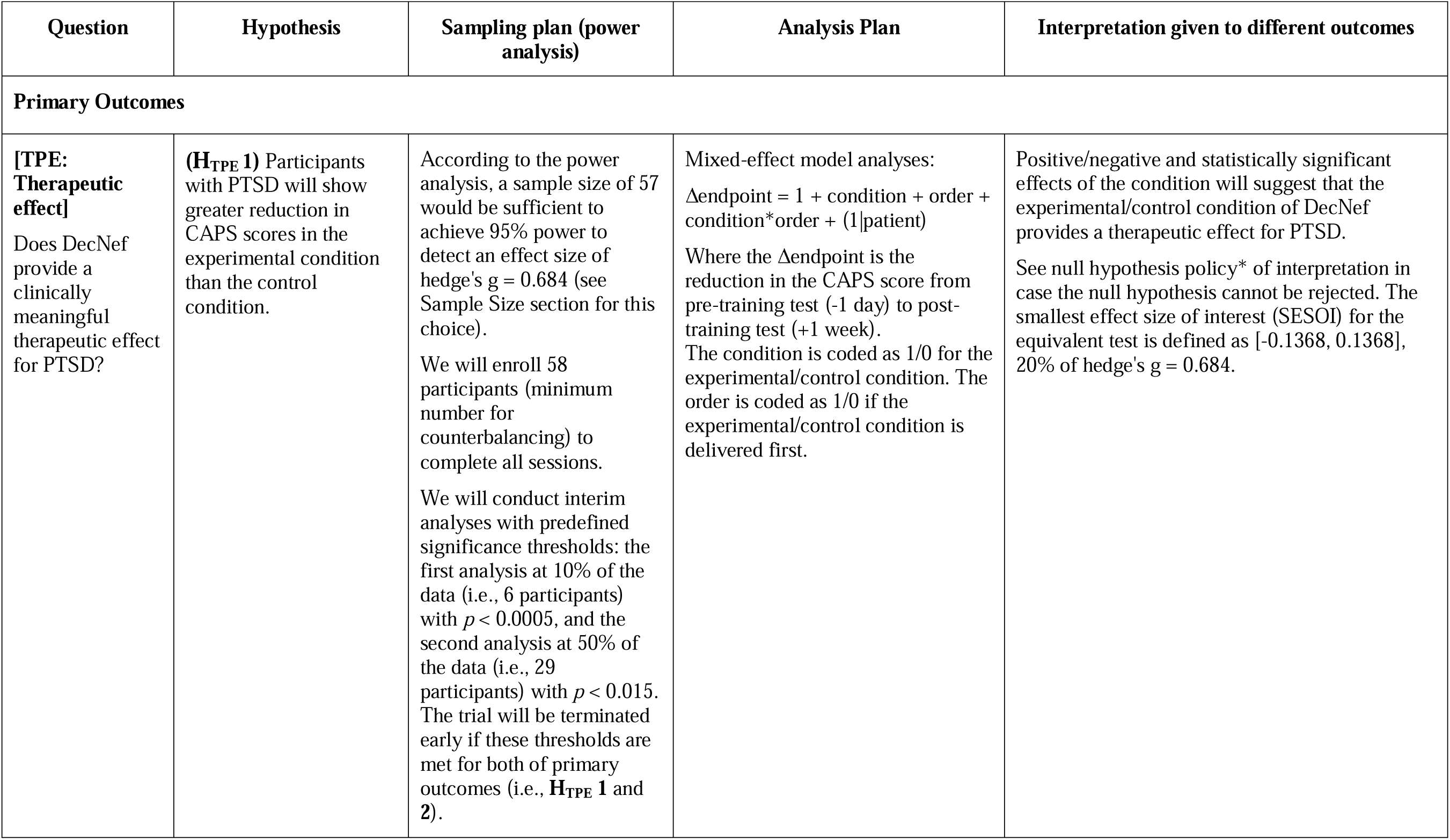
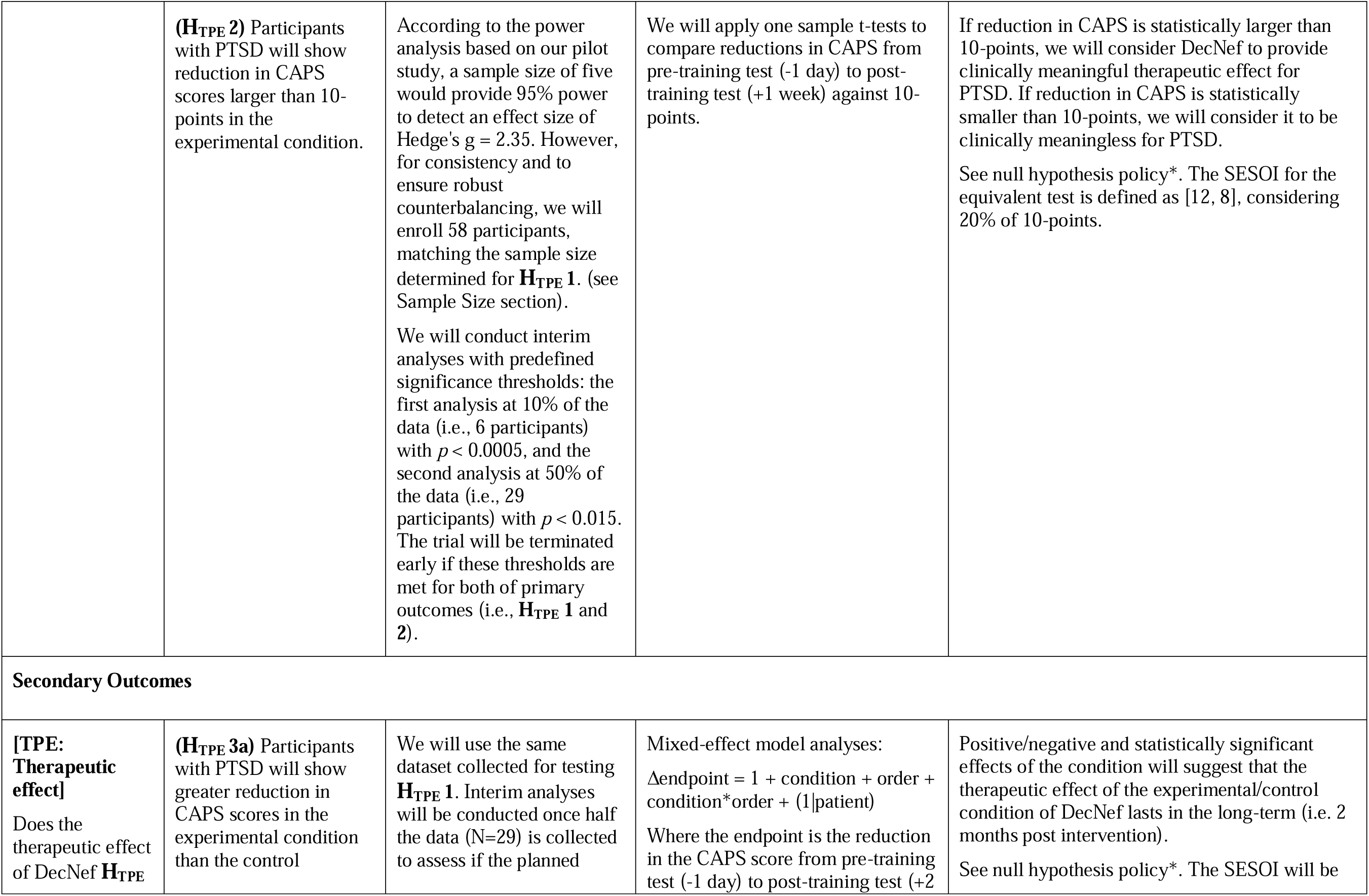
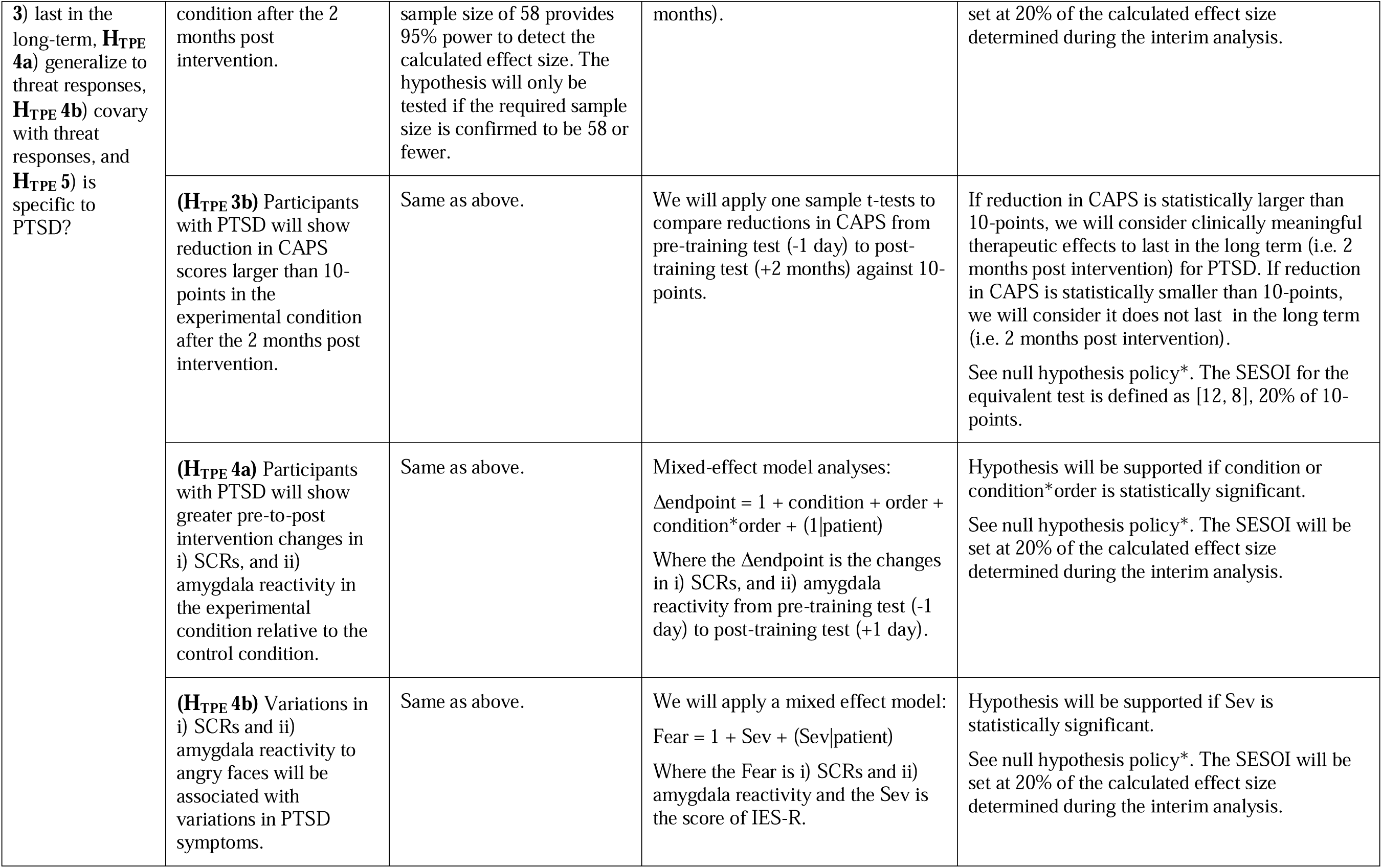
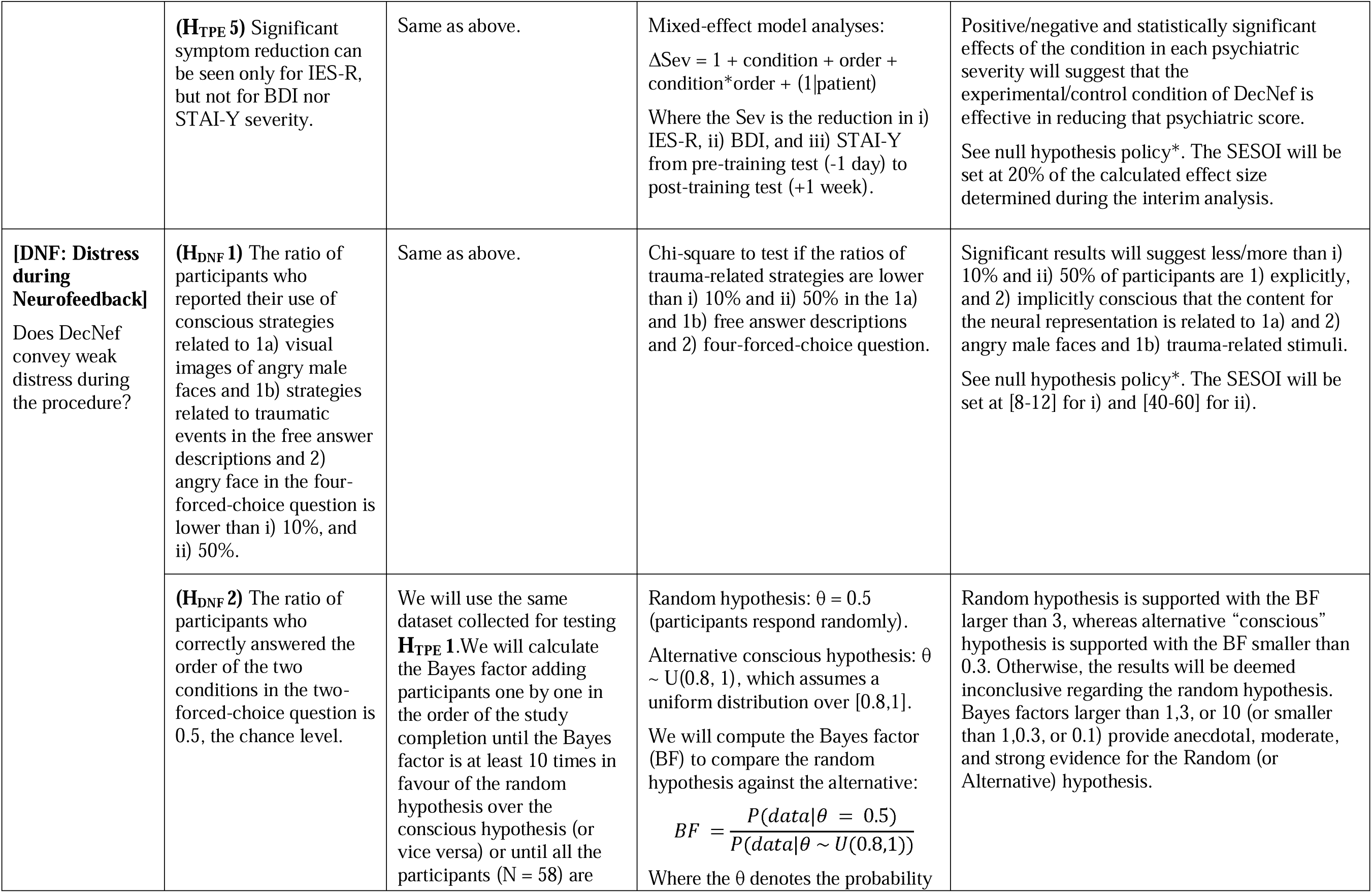
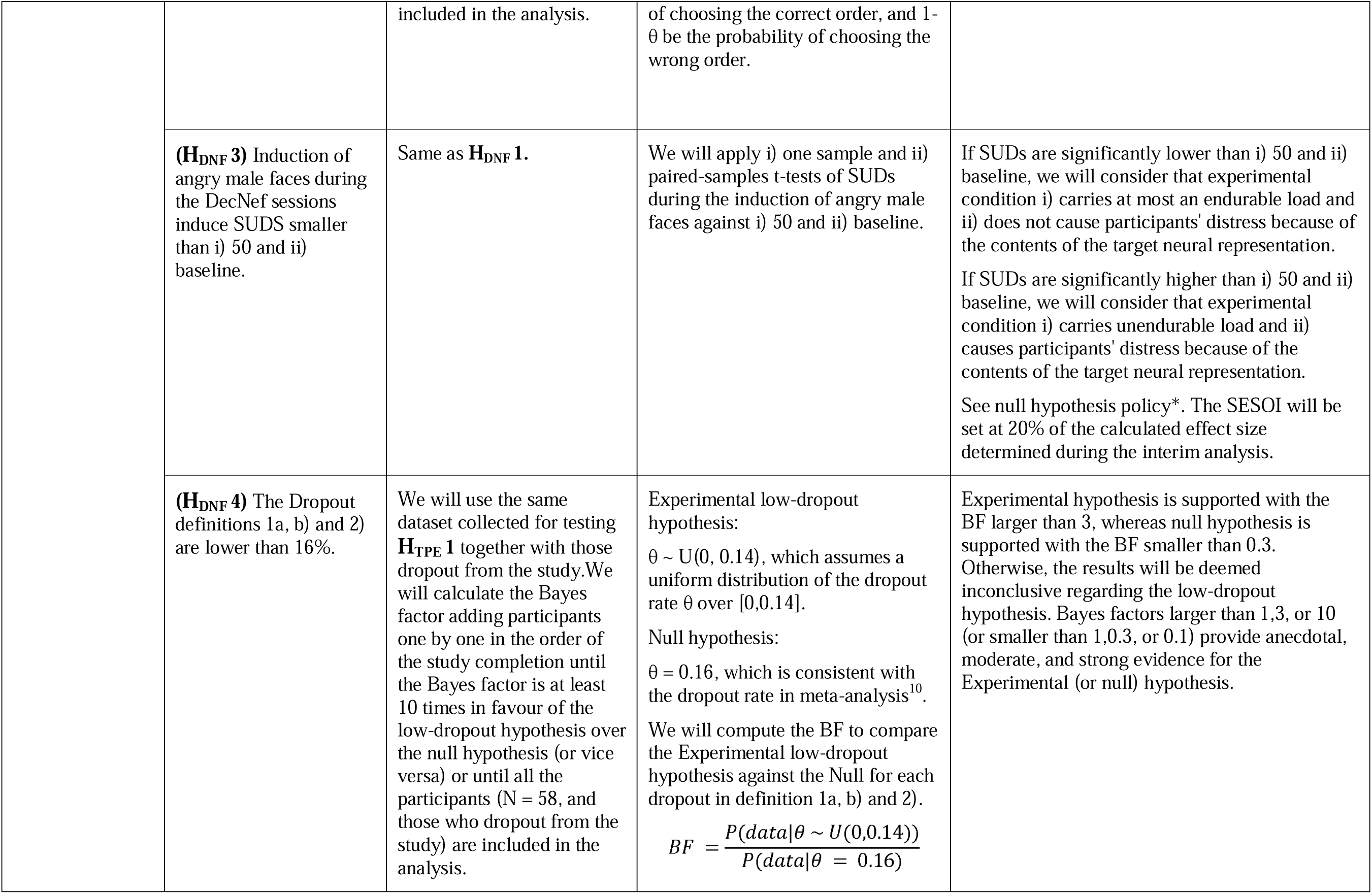
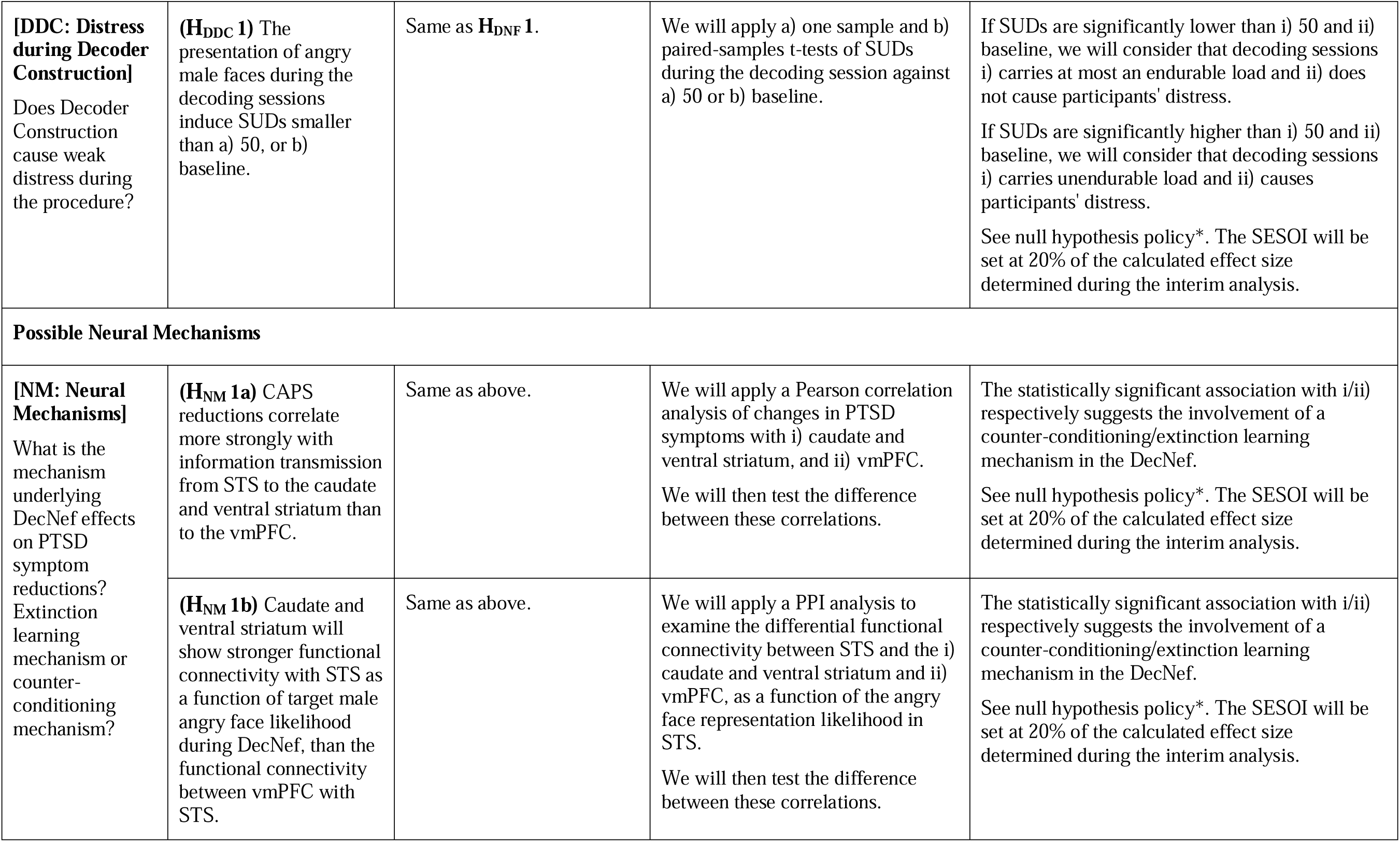
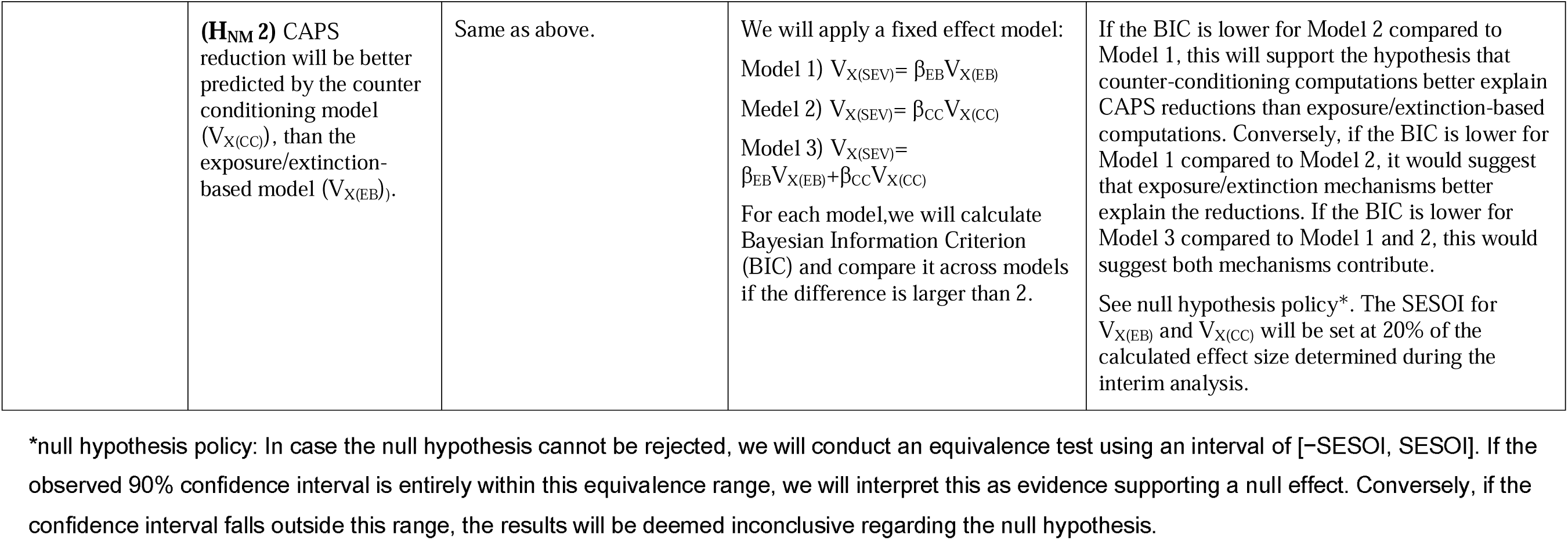
Design Table.

##### H_TPE_ 2 (Primary Outcomes)

**H_TPE_ 2** evaluates the DecNef effect using a clinically meaningful SESOI, defined as a 10-point reduction in CAPS (equivalent to hedge’s g = 2.35 from our pilot study). A two□tailed one-sample t-test (α = .05, β = .95) estimates that five participants are required to detect significance. Considering power analysis for **H_TPE_ 1**, the total sample size was set at 58 participants.

##### Other Hypotheses

For secondary analyses on therapeutic effects (**H_TPE_ 3-5**), distress during DecNef (**H_DNF_**) and during preparatory decoder construction (**H_DDC_**), and neural mechanisms (**H_NM_**) where effect sizes are uncertain, the dataset collected for primary outcomes (i.e. **H_TPE_ 1, 2**) will be repurposed. Interim analyses will be conducted at the halfway point (N=29) to assess whether a sample size of 58 provides 95% power for each hypothesis. Hypotheses will be tested only if the required sample size remains within this threshold. The SESOI for the equivalent test is defined as 20% of the interim-calculated effect size to ensure meaningful and robust hypothesis testing^69^. For the hypothesis where the Bayesian approach is adopted (**H_DNF_ 2** and **H_DNF_ 4**), we will calculate the Bayes factor adding participants one by one in the order of the study completion until the Bayes factor is at least 10 times in favour of the experimental hypothesis over the random hypothesis (or vice versa) or until all the participants (**H_DNF_ 2:** N = 58 that defined for **H_TPE_ 1; H_DNF_ 4:** N=58, plus all participants who dropped out of the study) are included in the analysis.

#### Inclusion criteria

PTSD patients will be recruited from Osaka Medical and Pharmaceutical University, the Seven-Mental Clinic (Osaka, Japan), and the Flower of Light Clinic for Mind and Body (Tokyo, Japan). Patients will be registered in this study by medical doctors, based on inclusion and exclusion criteria approved by Osaka Medical and Pharmaceutical University. Inclusion criteria require participants to be female, 20 to 55 years of age, meeting DSM-IV criteria for current PTSD diagnosis, having a score of ⍰45 on the clinician-administered PTSD scale (CAPS^25^), and having strong fear when passively viewing pictures of angry male faces, with a score of >60 on the SUDS. Participants will be also required to agree not to receive other psychotherapy for PTSD during the study treatment. Additionally, if participants are being treated with psychoactive medication, they must have been on a stable regimen (with no changes in drugs or doses) for at least 2 months prior to the DecNef session. Psychoactive medication can be modified after the post-training test (+2 months) of the first DecNef session block. However, the pre-training test of the second DecNef session block will be postponed until 2 months have passed on a stable regimen.

#### Exclusion criteria

Exclusion criteria are: active suicidality, a current diagnosis of substance dependence or psychosis, a history of moderate or severe head injury, and/or general contraindication to MRI. Participants with excessive head-motion during fMRI scanning (above 1.5 voxels on average), as well as those who provide fMRI data with lower classification accuracy (<56%) will be also excluded from analyses. We will use 56% as the cutoff, since pilot participants showed decoding accuracy higher than 56%, which led to clinically significant symptom improvement through DecNef in all participants.

### fMRI Analysis

#### Decoder construction

During decoder construction, for each trial, functional data will first be realigned to match coordinates of the first EPI image acquired in the Decoding session. Data will then undergo head-movement correction (using the realignment function of SPM12). We will use a gray-matter mask to extract fMRI data only from gray-matter voxels. Coordinates of the AAL atlas definition of the STS (Temporal_Sup_L, Temporal_Sup_R) will be extracted from this gray-matter using WFU_Pickatlas in MNI space and will then be retransformed to native space. Using the resulting coordinates in native space, we will extract the time-courses of BOLD signal intensities from the STS. Voxels with exceptional value will be removed. These will include voxels with exceptionally low BOLD signal intensities (mean <100) or those with exceptionally large variances (SD > 8 or SD <-8). We will remove a linear trend from the time-course. To further minimize baseline differences across runs, the time-course will be z-score normalized for each voxel in each run. We will extract pre-processed fMRI signals from the 6 s time-period of each trial of the Decoding session in which the face stimulus is constantly present on-screen, with a delay of 6 s to account for the hemodynamic delay. STS signals from this 6 s period, i.e., three TRs, will be averaged for each voxel. These will then be used to construct a decoder to classify activation patterns for angry male versus happy female faces. We will use Sparse logistic regression (SLR)^70^ to automatically select relevant voxels for classification. To estimate the validity of the constructed decoder, leave-one-run-out cross-validation will be performed. To be used in the DecNef session, we will train the decoder, without cross-validation, using data from all fMRI runs, i.e., 176 data points from 176 trials. Details on fMRI analysis tools are described in Supplementary Method.

#### Online fMRI analyses during DecNef

For online fMRI analyses during DecNef sessions, measured functional images will first undergo three-dimensional motion correction in real time so that they match coordinates of the first EPI image acquired in the Decoding session (using the realignment function of SPM12). Second, we will extract time-courses of BOLD signal intensities from each STS voxel that is relevant for decoding. Third, we will remove a linear trend from the time course. The BOLD signal time course will be z-score-normalized for each voxel using BOLD signal intensities measured for the initial 20 s fixation period of each run, after removing the initial 10 s from the whole 30 s fixation period. (Also see *DecNef session*). Fourth, we will create a data sample to calculate feedback by averaging BOLD signal intensities of each voxel for three TRs in the induction period, allowing for 6 s (three TRs) of hemodynamic delay. Before calculating feedback, the correlation between current and decoder activation patterns will be calculated to remove “error trials.” If the correlation is below 0.9, feedback will be presented as an error (a capital letter “E” will be displayed on the display). These trials will be defined as error trials. If the correlation is at or above 0.9, we will calculate the likelihood that neural activity from the induction period represents the target stimulus by applying the to-be-constructed decoder for the to-be-acquired data sample. The radius of the feedback disc will be proportional to the likelihood of target facial characteristics (angry male/happy female) assigned to each condition (experimental/control) on a given DecNef session block by the STS.

All of these procedures are consonant with previous DecNef studies^13,14,20^, except that we will use a pipeline based on SPM software (https://bicr.atr.jp/decnefpro/software/). This is in contrast to previous DecNef studies, as well as our pilot study, which relied on TurboBrainvoyager for head-motion correction.

#### Offline fMRI analyses for test sessions

Amygdala reactivity in response to angry male or happy female faces from the pre/post-training test session will be assessed for each test (pre-training, post-training (+1 day), post-training (+1 week), and post-training (+2 months), for each condition) for each participant based on subsequent offline analyses. An exemplar analysis workflow with FSL tools will be executed. First level analysis will utilize FILM (FMRIB’s Improved Linear Model) to set up a standard generalized linear model (GLM). Specifically, three types of GLM analyses will be performed for each voxel for each participant to extract contrast statistics between: 1) angry male versus happy female faces, 2) angry faces versus fixation, and 3) happy female faces versus fixation. By averaging the aforementioned contrast statistics (normalized coefficient, beta) calculated from voxels in the anatomically defined amygdala, we will define amygdala reactivity in response to 1) angry male versus happy female faces, 2) angry faces, and 3) happy female faces, respectively.

Amygdala reactivity in response to angry male or happy female faces from the decoding session will be assessed for each participant in a similar manner.

### Skin conductance responses Analysis

Consistent with a previous study^17,20^, stimulus-elicited SCRs for each test to be analyzed, i.e., a pre-training test or a post-training test, consisting of two fMRI runs, will be defined as averaged SCRs for happy female faces subtracted from those for angry male faces. We will use the following methods of SCR extraction: 1) Before analyses, a band-pass filter (transmission range, 0.05-1 Hz) will be applied to each fMRI run to remove noise from the data; 2) The SCR for each stimulus presentation will be calculated as the maximum SCR that occurs during stimulus presentation (from 0.5 s after onset of the presentation until the end of the presentation) as compared to baseline, which will be defined as the averaged skin conductance from 2 s before stimulus presentation; 3) SCRs lower than 0.02 μSiemens will be scored as 0; 4) SCRs will be square root transformed to correct for the skewness of their distributions^17^; 5) SCRs from each test will then be averaged separately for angry male faces and for happy female faces, after exclusion of the initial dummy trial of each fMRI run to remove irrelevant orienting effects (see pre/post-training test session).

### Analytical Plan

Analyses will be performed only for data of completers. In cases where the null hypothesis cannot be rejected, we will perform an equivalence test to determine if the observed 90% confidence interval falls entirely within the equivalence range. The equivalence range will be defined as ±20% of the effect size used in the power analysis for each hypothesis^69^.

To address potential biases from randomization and dropouts, supplementary analyses will include baseline comparisons (**SH_BC_ 1 & 2**) and intent-to-treat analyses (**SH_TPE_ 1-3**). Baseline comparisons will evaluate equivalence of demographic and clinical characteristics between completers and dropouts as well as across randomized groups. Intent-to-treat analyses will replicate the main analyses, incorporating data from all participants, including those who dropped out.

Neyman-Pearson inference will test the alternative hypothesis (predicting a difference), while Bayesian hypothesis testing will assess the random hypothesis (predicting no difference: **H_DNF_ 2**) and compare the two distributions (**H_DNF_ 4**).

#### Analysis of PTSD severity (H_TPE_ 1, 2, and 3)

##### H_TPE_ 1

We will analyze the main effect of DecNef conditions, either experimental or control, on potential reductions in CAPS scores (ΔSev) using the following mixed-effect model:

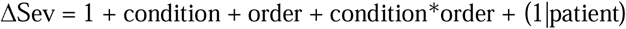

where ΔSev will be defined for the post-training test (+1 week) as a reduction from the pre-training test (−1 day). The condition denotes the type of condition (experimental vs control condition) while the order denotes the order of the condition.

##### H_TPE_ 2

We will analyze if the DecNef effect exceeds a clinically defined meaningful SESOI, defined as a 10-point reduction. Specifically, we will apply one sample t-tests of reduction in CAPS from pre-training test (−1 day) to post-training test (+1 week) against 10-point reduction.

##### H_TPE_ 3a & b

We will conduct a similar analysis for **H_TPE_ 1** & **2** using a post-training test (+2 months) instead of post-training test (+1 week).

##### Supplementary Hypotheses (see Supplementary *Design Table*)

We will conduct a similar analysis for **H_TPE_ 1-3** in an intent-to-treat analyses instead of complete analyses (**SH_TPE_ 1-3**).

#### Analysis of physiological threat responses (H_TPE_ 4)

##### H_TPE_ 4a

The mean SCRs/amygdala reactivity for angry male faces minus those for happy female faces will be analyzed using the mixed-effects model for **H_TPE_ 1**, the CAPS analyses. Baseline correction using the reactivity to fear-irrelevant stimuli—specifically, happy female faces—enables the evaluation of fear-specific reactivity^17,20,71^. In contrast to CAPS analyses, we will use data from post-training tests (+1 day) instead of post-training tests (+1 week).

##### H_TPE_ 4b

Whether variations in PTSD severity as measured using IES-R (Sev) are accompanied by variations in SCRs/amygdala reactivity (Fear) will be analyzed using the following mixed effect model:

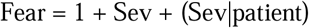

Here, Fear and Sev will be derived from all assessment time points: pre-training test (−1 day) and post-training tests (+1 day, +1 week, +2 months).

##### Supplementary Hypotheses (see Supplementary *Design Table*)

Extracted SCRs/amygdala reactivity for each face type (angry male and happy female) will also be analyzed in a similar vein to **H_TPE_ 4a.** The analyses explore the possibility that reactivity for a specific face type, including happy female faces, may be affected by DecNef, considering that happy female faces are reinforced in the control DecNef condition (**SH_TPE_ 4a**).

#### Analysis of questionnaire-based psychological assessments (H_TPE_ 5)

##### H_TPE_ 5

The reduction of IES-R, BDI, and STAI-Y scores will be analyzed separately, but in a similar manner to CAPS analyses (see analysis for **H_TPE_ 1**). We will use the IES-R, a self-report questionnaire instead of the CAPS, a structured interview, to enable fair comparison with other self-report measures such as the BDI and the STAI-Y.

##### Supplementary Hypotheses (see Supplementary *Design Table*)

If **H_TPE_ 1** demonstrates the efficacy of DecNef, supplementary analyses will investigate its effects on specific PTSD symptom clusters (**SH_TPE_ 5**), examine how PTSD subtypes influence therapeutic outcomes (**SH_TPE_ 6**), and explore detailed associations between physiological threat responses and symptom characteristics (**SH_TPE_ 7**).

#### Evaluating consciousness, distress, and dropout with DecNef (H_DNF_)

##### H_DNF_ 1

Conscious awareness of neural representations of trauma-related stimuli, i.e., angry male faces, during DecNef will be assessed as follows: We will first evaluate ratios of participants who reported their use of conscious strategies related to 1) visual images of angry male faces and 2) strategies related to traumatic events, including visual images of angry male faces, based on their open-ended responses, at the end of each DecNef session (see DecNef session). Categorization of strategy will be performed independently by two judges, including the doctor-in-charge. The corresponding author will categorize the strategy in case of conflict. We will use a chi-square test to examine whether ratios of conscious strategies are lower than i) 10% and ii) 50%. If they are lower than these thresholds, we will consider that explicit consciousness, a form of side effect, i) are not very frequent^72^, and ii) do not emerge in more than half of the patients. Further, chi-square tests will be performed on the four-forced-choice question asking participants to report the content of induced neural representations during DecNef. Specifically, we will examine if the ratios of participants who selected “an angry man” are lower than ii) 10% and ii) 50%. If they are lower than these thresholds, we will consider that implicit consciousness, a form of side effect, i) are not very frequent^72^, and ii) does not emerge in more than half of the patients. Chi-square tests will be replaced with Fisher’s exact test if expected frequencies in cells are below 5.

##### H_DNF_ 2

To assess differential conscious experiences beyond H_DNF_ 1, we will test whether participants can identify the condition order after the experiment, knowing that the experimental condition involves trauma-related content. Accurate reporting would indicate conscious awareness during DecNef, undermining its clinical benefits. Since we predict no such awareness, we will compare the Random and Alternative Conscious hypotheses using Bayesian testing.

The Random hypothesis assumes random guessing (θ = 0.5). Where the θ denotes the probability of choosing the correct order, and 1-θ be the probability of choosing the wrong order. The Alternative Conscious hypothesis posits awareness sufficient for accurate reporting (θ ∼ U(0.8, 1)), modeled as a uniform distribution over [0.8,1], allowing for 20% variation as in SESOI estimation. Here, we did not consider awareness that is insufficient for accurate reporting but still above chance (e.g. θ = 0.6), as such vague awareness would not necessarily undermine DecNef’s clinical benefits.

##### H_DNF_ 3

Distress during DecNef will be assessed with subjective measures of SUDS levels. One-sample/paired-samples t-test will be performed to assess whether the averaged SUDS during the experimental condition is lower than a) 50 (the strongest load that is still considered endurable), b) the baseline score measured before the first fMRI of each DecNef session. If SUDS during the DecNef is <50, we will consider that experimental condition carries at most an endurable load. If SUDS is lower than baseline, we will consider that the experimental condition does not enhance participant distress because of the contents of the target neural representation.

##### H_DNF_ 4

We will examine if the dropout rate in our study follows a distribution lower than the pooled dropout rate reported in the meta-analysis of RCTs of PTSD psychotherapy in general (mean = 16%, 95% CI: [14-18%])^10^. Specifically, we will compare the Null and Experimental hypotheses using Bayesian testing. The Null hypothesis assumes a dropout rate θ equivalent to the meta-analysis (θ = 0.16)^10^. The Experimental hypothesis posits lower distribution (θ ∼ U(0, 0.14)), modeled as a uniform distribution over [0,0.14]. In both hypotheses, the number of dropouts follows a binomial distribution as a function of θ (X ∼ B (N, θ)). In the Bayesian testing, the evidence for the experimental hypothesis will be calculated using a Beta distribution Beta (1,1), rescaled over the interval [0, 0.14].

Three types of dropout (1-a, 1-b, and 2) will be defined, as follows:

1) Dropout at any point in the entire procedure, excluding genuine interval period:

1-a) Dropout for any reason
1-b) Dropout upon request of the participant or the doctor-in-charge (excluding reasons unrelated to participant health, such as MRI malfunction, below-chance-level decoding accuracy, excessive head movements in the scanners, family concerns, or traffic conditions).
2) dropout during one of the three consecutive days for both DecNef sessions.

The dropout rate in each definition will be defined as the percentage of total dropouts relative to all participants entering the study. Since the RCTs in the meta-analysis did not adopt a crossover design and therefore did not include an interval period, dropout definition 1) will not include participants who left the study during the genuine interval period, from post-training test 1 (+2 months) to pre-training test 2 (−1day). Dropout definition 1-a) is the most conservative definition. If this dropout rate is lower than 14-18%, we will consider DecNef superior to current psychotherapies in terms of therapy design. The dropout definition 1-b) provides a fair measure for comparison with current psychotherapies. If this dropout rate is lower than 14-18%, we will consider DecNef superior to current psychotherapies, at least in terms of therapy adherence. The dropout definition 2) provides insights into the tolerability of DecNef in comparison with current psychotherapies including prolonged exposure.

##### Supplementary Hypotheses (see Supplementary *Design Table*)

We will conduct supplementary analyses to determine whether experimental and control conditions of DecNef produce differing conscious experiences. These assessments will examine distress levels (**SH_DNF_ 1**), dropout rates (**SH_DNF_ 2**), and the use of conscious strategies (**SH_DNF_ 3**) during DecNef. We will also compare physiological threat responses (**SH_DNF_ 4**) across conditions.

#### Evaluating consciousness and distress during decoder construction (H_DDC_)

##### H_DDC_ 1

To evaluate distress during decoder construction, one sample/paired-samples t-test will be performed to assess whether the averaged SUDS during the decoding session is lower than 1) 50 (the strongest load that is still considered endurable), and 2) the baseline score that will be measured before the first fMRI session for the decoding in a similar manner to the SUDs analyses during neurofeedback.

##### Supplementary Hypotheses (see Supplementary *Design Table*)

We will conduct supplementary analyses to determine whether presentations of trauma-related angry male faces during decoder construction produce different conscious experiences (**SH_DDC_ 1**) and physiological threat responses (**SH_DDC_ 2**) from those produced by presentations of happy female faces.

#### Neural mechanisms of DecNef effects (H_NM_)

##### H_NM_ 1a

The aim of this analysis is to test whether DecNef’s underlying mechanisms map onto extinction learning or counter-conditioning. To do so, we will examine whether core regions in extinction learning, vmPFC^59,73^, and counter-conditioning, caudate and ventral striatum^74,75^, are engaged when the angry face pattern is induced in STS during DecNef. To estimate engagement of these areas, we will calculate their relative information transmission with the STS. More specifically, we will calculate the degree to which their activity patterns predict the angry face likelihood in STS activity patterns^11,13^. To this end, we will train a SLR decoder^70^ so that activation patterns in each ROI reconstruct the STS likelihood. Here, the likelihood in STS will be identical to the likelihood to-be-fed back to participants during DecNef. Leave one-run out cross-validation will be performed to estimate the validity of the constructed decoder. For each ROI, we will analyze the across-participant Pearson correlation of symptoms reduction (pre-training test versus post-training test (+1 week)) with information transmission. We will then test the difference between these correlations. If DecNef shares its mechanism with exposure therapy, then vmPFC should be positively correlated with symptom reduction. If DecNef shares its mechanism with counter-conditioning, then information transmission in caudate and ventral striatum should be positively correlated with symptom reduction.

To comprehensively examine information transmission in a data-driven manner, we will perform a whole-brain MVPA using a searchlight method, as described in the Supplementary methods. Anticipating the interests of the general audience, we will conduct supplementary analyses with the amygdala, as well as caudate and ventral striatum separated.

##### H_NM_ 1b

To illuminate mechanisms underlying DecNef, we will further examine how angry face representations induced in STS may lead to changes in the STS connectivity with other brain areas. Specifically, we expect fear/reward circuits to interact with the STS during the successive DecNef trials if DecNef is involved in fear extinction/reinforcement learning. We will conduct a psychophysiological interaction (PPI) analysis^76^ to examine the differential functional connectivity of the STS with other brain regions, as a function of the angry face representation likelihood in the STS, i.e., psychological variable, for each ROI, i.e., vmPFC, and caudate and ventral striatum, as well as for each voxel in the whole brain. To this end, we will test the difference between these correlations.

The GLM model in the PPI analysis will include regressors for the angry face representation likelihood (psychological variable), the z-normalized time course of the seed ROI, and the PPI term (seed time course x likelihood). We will code the psychological variable as 1 or −1 to cover the induction and rest periods of trials (total 12 s) with high (> 50%) versus low (< 50%) likelihood (the disc size reflecting the induction likelihood) respectively, convolved with a canonical HRF. Additional regressors of no interest will be the initial rest period (30 s), the feedback period, and the fixation period (each coded separately for high and low likelihood trials), as well as six motion parameters, the day of DecNef sessions, and the experimental run number. We will use voxels within the STS that are activated with a liberal threshold (uncorrected P<.1) during the induction period (irrespective of angry face representation likelihood) relative to the fixation period. We will conduct these analytical steps within naive coordinates of each participant. Then, in the MNI space, we will conduct a group level analysis to obtain a map of voxels showing significant differential connectivity with the STS as a function of angry male face representation likelihood (P<0.05; corrected with permutation procedure^77^).

In a similar vein to **H_NM_ 1a,** we will conduct supplementary analyses with the amygdala, as well as caudate and ventral striatum separated.

##### T_NM_ 2

There are two possible learning processes underlying the fear-reduction effects of DecNef. One is through effects of exposure, where neural representations of feared stimuli are induced without actual fear or distress. The other is through effects of counter-conditioning, where such representations are paired with reward. In order to dissociate potential effects of exposure from those of counter-conditioning, we will apply a mathematical model, as in our previous study^22^. The model dissociates DecNef effects derived from exposure therapy (*V_X_*_(*EB*)_) and counter-conditioning (*V_X_*_(*CC*)_) separately, on the basis of the Rescorla-Wagner model and synaptic plasticity rules. Detailed information can be found elsewhere^22^. Briefly, (*V_X_*_(*EB*)_) is assumed to be linearly proportional to the number of “successful” trials, where the likelihood for the target activity pattern is above chance. (*V_X_*_(*CC*)_) of each trial is also assumed to be proportional to the induction likelihood of the target pattern multiplied by the amount of the reward that the participant obtains in the trial. Finally, we assume that the DecNef effect is caused by the weighted linear summation of (*V_X_*_(*EB*)_) and (*V_X_*_(*CC*)_).

Based on these assumptions, the total effect is given as follows:

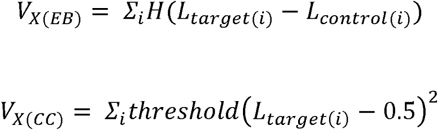

Then, we will compare following three models:

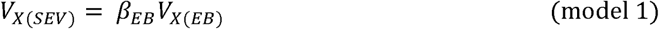

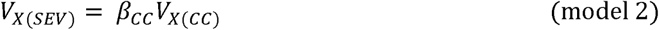

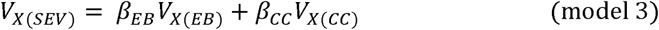

where *V_X_*_(*SEV*)_ is symptom reduction from the pre- to post-training test. The β_EB_ and β_CC_ are the coefficients of *V_X_*_(*EB*)_ and *V_X_*_(*CC*)_, respectively. H(X) is the Heaviside step function, which is 1 if X > 0 and 0 otherwise. The threshold(X) = X if X > 0, and 0 otherwise. We will test these models using fixed effect models. The model comparison will be conducted using Bayesian Information Criterion (BIC).

## Data availability

All data and materials will be made publicly available.

## Code availability

All analysis code will be made publicly available.

## Acknowledgements

This study is funded by the Innovative Science and Technology Initiative for Security (JPJ004596), ATLA, AMED (JP20dm0307008), and KDDI collaborative research contract. The funders had no role in the study design, data collection and analysis, the decision to publish, or in preparation of the manuscript.

We thank Kaori Nakamura and Chika Hosomi for their help with scheduling and conducting the pilot experiment.

## Author contributions

TC, HL, MK, and AK conceived the research. TC, KI, JET, AC, TK, KA, HL, MK, AK, and TK designed the research. TC developed the decoding and DecNef task in consultation with AC, TK, KA, BS, and AK. TC analyzed the pilot data in consultation with KA, AK, and MK. TC conducted the power analysis in consultation with KA, AK, and MK. TC, IK, KA, HL, MK, AK, and KT developed the analysis plan. TC and JET prepared the manuscript with feedback from all co-authors. TC, KI, JET, SB, HT, MS, and TK coordinated the implementation of the project. TC, KI, JET, KN, SB, HT, MS, and TK will contribute to data collection. All co-authors will review and approve the final manuscript.

