## Supplementary information for "PTSD therapy with fMRI-decoded neurofeedback bypassing conscious exposure: a randomized, double-blind, placebo-controlled study"

**Sections of a Stage 1 Registered Report**

^1^ The Department of Decoded Neurofeedback, Computational Neuroscience Laboratories, Advanced Telecommunications Research Institute International, Kyoto, Japan, ^2^ The Department of Psychiatry, Self-Defense Forces Hanshin Hospital, Kawanishi, Japan, ^3^ Seven-Mental Clinic, Osaka, Japan, ^4^ Flower of Light Clinic for Mind and Body, Tokyo, Japan, ^5^ Graduate School of Science and Technology, Nara Institute of Science and Technology, Nara, Japan, ^6^ The Department of Neuropsychiatry, Osaka Medical and Pharmaceutical University, Osaka, Japan, ^7^ Department of Neuropsychiatry, Kumamoto University Faculty of Life Sciences, Kumamoto, Japan, ^8^ The Department of Psychiatry, National Defense Medical College, Tokorozawa, Japan, ^9^ Graduate School of Information Science and Technology, The University of Tokyo, Tokyo, Japan, ^10^ RIKEN Center for Brain Science, Wako, Japan, ^11^ XNef, Kyoto, Japan, ^12^ Department of Engineering Science, Institute of Biomedical Engineering, University of Oxford, Old Road Campus Research Building, Oxford, UK, ^13^ The Department of Neural Computation for Decision-making, Cognitive Mechanism Laboratories, Advanced Telecommunications Research Institute International, Kyoto, Japan, ^14^ Sony Computer Science Laboratories, Inc., Tokyo, Japan, ^15^ The Department of Neuropsychiatry, Osaka Medical and Pharmaceutical University, Osaka, Japan

**Supplementary information**

*Pilot study*

**Overview**

Seven patients with PTSD were enrolled (Supplementary Table 1). We first performed DecNef of experimental conditions on five participants with PTSD. After confirming that PTSD severities were reduced from pre-test to post-test in those participants, two of the initial five participants subsequently underwent a control condition, whereas the remaining three participants did not. Two additional participants also underwent a control condition, and only one of them underwent an experimental condition afterward, while the other did not. To summarize, two participants underwent an experimental condition followed by a control condition. One participant underwent a control condition followed by an experimental condition. Three participants underwent only the experimental conditions, and one patient participated only in control conditions. Three participants who underwent both conditions had an interval of more than 6 months between the two conditions. These pilot studies were performed in single-blinded fashion as opposed to double-blinded fashion in this pre-registered study. That is, although experimenters were aware of which condition participants were undergoing, participants were unaware of which condition they were experiencing. In addition, analyses of fMRI data differed in the pilot study from the proposed study. Specifically, we used mrVISTA software (<http://vistalab.stanford.edu/software/>) in pre-processing for the decoding analysis and Turbo BrainVoyager software (Brain innovation) in the DecNef online analysis. Other procedures of the pilot study were consistent with the proposed study.

**Supplementary Table 1. Procedures in the pilot study**

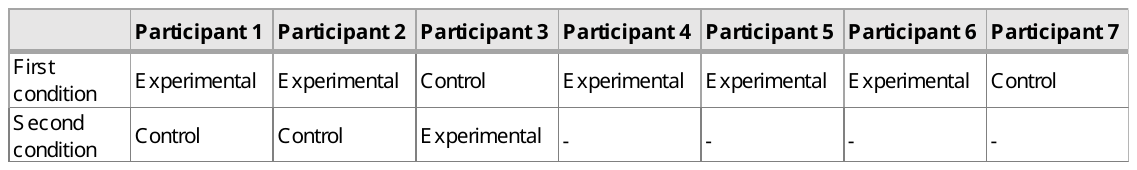

**Preprocessing of fMRI data using mrVista**

fMRI signals were preprocessed with mrVista software, developed at Stanford University (<http://vistalab.stanford.edu/software/>). We first conducted 3D motion correction without spatial and temporal smoothing for functional images. We then conducted rigid-body transformations to align the images to the structural image for each participant. We extracted fMRI signals from only the gray matter using a gray matter mask. The superior temporal sulcus (STS) ROI was defined anatomically with freesurfer segmentation (https://surfer.nmr.mgh.harvard.edu/).

*Supplementary method*

**Semi-randomization procedures**

In the main experiment, the order of experimental and control conditions will be semi-randomized using the following procedures. First, participants will be classified into moderate PTSD (45 ≤ CAPS ≤ 60; M-PTSD) or severe PTSD (CAPS > 60) groups based on their CAPS score at the time of enrollment. Then, participants will be assigned a condition sequence (experimental condition first or control condition first) to balance the proportion of participants in the moderate and severe PTSD groups. Specifically, a patient from the moderate (or severe) PTSD group will be assigned to a condition sequence with a smaller number of participants from the same moderate (or severe) PTSD group. In case the number of participants from the moderate (or severe) group is identical in the two condition sequences, participants will be assigned to the condition sequence with a smaller total number of participants (from moderate and severe groups combined). If the number of participants in both severity groups is equal, the condition sequence of the next participant will be assigned randomly. These procedures will be continued until 58 participants are enrolled. In case of dropout, we will continue recruiting participants until 58 participants complete the whole procedure. The order of conditions will be assigned randomly to these additional participants regardless of their PTSD scores, so as to complete the experiment with equal patient numbers in both condition sequences. Those procedures are automated with Matlab 2020b scripts to maintain the blindness of the operators to the condition assignments.

**fMRI analysis tools**

Offline analyses will be performed using fMRIprep^1^ 1.4.1 (RRID:SCR_016216), which is based on Nipype 1.1.6 (RRID:SCR_002502). fMRIprep is a robust pipeline that can also be used for task-based functional MRI data^2^. For multivariate information transmission analysis with whole brain searchlight analysis^3^ and psychophysiological interaction (PPI) analyses^4^, we will use the Nilearn 0.9.2 toolbox^5^ and the Sparse logistic regression (SLR) toolbox^6^.

Online analyses will be performed using Statistical Parametric Mapping (SPM12; Welcome Department of Imaging Neuroscience, London, UK) with minimal preprocessing to achieve real-time feedback. Since SPM software is the most widely used for fMRI analyses, this will enhance application of DecNef in other laboratories, as well as in clinical settings. SPM and WFU_Pickatlas (https://www.nitrc.org/projects/wfu_pickatlas), an extension toolbox, will be used in Matlab 2020b. Although fMRI data from the decoding session will be analyzed offline, they will be pre-processed as in online analysis. This will make decoder results compatible with those from online analyses conducted during DecNef sessions. For decoding, SLR toolbox^6^ will be used. For DecNef, DecNef toolbox (<https://bicr.atr.jp/decnefpro/software/>) will be used.

**“Information Transmission” in a whole-brain MVPA analysis**

We will conduct a whole-brain MVPA analysis with a searchlight method^3^ to highlight voxels showing significant information transmission. We will create whole-brain ROIs by shifting a center of sphere ROI (radius=15 mm, M=266 voxels) to each voxel of the whole brain within the naive coordinates of each patient. For each sphere ROI, information transmission during DeNef will be calculated as described in the ROI analysis, which will yield a whole-brain map of the Fisher-transformed correlation coefficient, i.e., information transmission. We will then project the map of each patient to MNI coordinates and spatially smooth them (Gaussian filter FWHM=6 mm). For each voxel, we will perform a one-sample t-test on the Fisher-transformed correlation coefficient against 0, chance level. To adjust for the multiple comparison, we will extract voxels with significant information transmission with a permutation procedure^66^. We will iteratively calculate a t-value 1000 times based on a randomized data set. We will define the 5th percentile of the histogram of the t-values as a threshold for that voxel. We will consider any voxel with a t-value above the threshold as having significant information transmission.

*Supplementary analyses*

The supplementary analyses aim to deepen understanding of DecNef's effects, including baseline group comparisons (**SH_BC_ 1, 2**), addressing dropout biases via intent-to-treat analyses (**SH_TPE_ 1, 2, 3a, 3b**), examining nuanced changes in physiological threat responses to specific face types from pre-test to post-test (**SH_TPE_ 4a**), and evaluating DecNef effects on PTSD symptom clusters (**SH_TPE_  5**), PTSD subtypes (**SH_TPE_ 6**), and correlations between across-patient variabilities in symptom imbalance and physiological threat responses (**SH_TPE_ 7**). Consciousness and distress during DecNef (**SH_DNF_ 1-4**) and decoder construction (**SH_DDC_ 1, 2**) are also assessed using mixed models and chi-square tests. These analyses investigate distress, dropout, and physiological/neural responses, ensuring robustness of findings while addressing potential biases.

***Supplementary Table 2. Supplementary Design Table***

| **Question** | **Hypothesis** | **Sampling plan** | **Analysis Plan** | **Interpretation given to different outcomes** |
| --- | --- | --- | --- | --- |
| **[BC: Baseline comparison]**  Is the DecNef effect affected by group differences at the baseline? | **(SH_BC_ 1)** Participants assigned to different condition order groups have different baseline demographics. | We will use the same dataset collected for testing **H_TPE_ 1**.  Interim analyses will be conducted once half the data (N=29) is collected to calculate the smallest effect size of interest (SESOI). | ・We will apply independent-samples t-tests to compare 1) age and 2) baseline severity as measured by CAPS between groups.  ・We will apply a Chi-square test to compare the ratio of sex between groups. | The statistically significant results will suggest that the main results might be affected by the baseline differences between groups.  See null hypothesis policy*. The SESOI will be set at 20% of the calculated effect size determined during the interim analysis. |
|  | **(SH_BC_ 2)** Completers and dropouts have differential baseline demographics. | We will use the same dataset collected for testing **H_TPE_ 1** together with those who dropped out.  Interim analyses will be conducted once half the data (N=29) is collected to calculate the SESOI. | ・We will apply independent-samples t-tests to compare 1) age and 2) baseline severity as measured by CAPS between completers and dropouts.  ・We will apply a Chi-square test to compare the ratio of sex between groups. | The statistically significant results will suggest that the main results might be affected by the baseline difference of completers from dropouts.  See null hypothesis policy*. The SESOI will be set at 20% of the calculated effect size determined during the interim analysis. |
| **[TPE: Therapeutic effect]**  Do therapeutic effects on PTSD symptoms hold in intent-to-treat samples, (**SH_TPE_ 1, 2, 3a, 3b**)? Are therapeutic effects on physiological threat responses specific to each face stimulus type (**SH_TPE_ 4a**)? Do therapeutic effects emerge in each PTSD symptoms cluster (**SH_TPE_ 5**)? Are therapeutic effects selective to a certain PTSD subtype (**SH_TPE_ 6**)? Is across-patient symptom imbalance variability correlated with that of physiological threat responses (**SH_TPE_ 7**)? | **(SH_TPE_ 1)** Participants with PTSD will show ~~1)~~ greater reduction in CAPS scores in the experimental condition than the control condition in the intent-to-treat samples including those who dropped out. | We will use the same dataset collected for testing **H_TPE_ 1** including data from participants who dropped out. This analysis will be conducted only when the hypothesis **H_TPE_ 1** yields statistically significant results.  The smallest effect size of interest (SESOI) is derived from **H_TPE_ 1**. | Mixed-effect model analyses:  Δendpoint = 1 + condition + order + condition*order + (1\|patient)  Where the Δendpoint is the reduction in the CAPS score from pre-training test (-1 day) to post-training test (+ 1 week). | The statistically significant effects of the condition in the same direction as the main results will suggest that the main results are not biased by dropout.  See null hypothesis policy* of interpretation in case the null hypothesis cannot be rejected. The smallest effect size of interest (SESOI) for the equivalent test is defined as [-0.1368, 0.1368], 20% of hedge's g =0.684 from H_TPE_ 1. |
|  | **(SH_TPE_ 2)** Participants with PTSD will show reduction in CAPS scores larger than 10-points in the experimental condition in the intent-to-treat samples. | We will use the same dataset collected for testing **H_TPE_ 2**. This analysis will be conducted only when the hypothesis **H_TPE_ 2** yields statistically significant results. | We will apply one sample t-tests of reduction in CAPS from pre-test (-1 day) to post-test (+ 1 week) against 10-point. | If reduction in CAPS is statistically larger than 10-points, we will consider that the main effects are not biased by dropout.  See null hypothesis policy*. The SESOI for the equivalent test is defined as [12, 8], 20% of 10-points. |
|  | **(SH_TPE_ 3a)** Participants with PTSD will show greater reduction in CAPS scores in the experimental condition than the control condition after the 2 months post intervention in the intent-to-treat samples. | We will use the same dataset collected for testing **H_TPE_ 3a**. This analysis will be conducted only when the hypothesis **H_TPE_ 3a** yields statistically significant results. | Mixed-effect model analyses:  Δendpoint = 1 + condition + order + condition*order + (1\|patient)  Where the Δendpoint is the reduction in the CAPS score from pre-training test (-1 day) to post-training test (+ 2 months). | The statistically significant effects of the condition in the same direction as the main results will suggest that the main results are not biased by dropout.  See null hypothesis policy*. The SESOI will be consistent with that of **H_TPE_ 3a**. |
|  | **(SH_TPE_ 3b)** Participants with PTSD will show reduction in CAPS scores larger than 10-points in the experimental condition after 2-months post intervention in the intent-to-treat samples. | We will use the same dataset collected for testing **H_TPE_ 3b**. This analysis will be conducted only when the hypothesis **H_TPE_ 3b** yields statistically significant results. | We will apply one sample t-tests of reduction in CAPS from pre-test (-1 day) to post-test (+ 2 months) against 10-point. | If reduction in CAPS is statistically larger than 10-points, we will consider that the main effects are not biased by dropout.  See null hypothesis policy*. The SESOI for the equivalent test is defined as [12, 8], 20% of 10-points. |
|  | **(SH_TPE_ 4a)** Participants with PTSD will show greater pre-to-post intervention changes in threat responses to 1) angry male faces or 2) happy female faces in the experimental condition than the control condition. | We will use the same dataset collected for testing **H_TPE_ 4a**.  Interim analyses will be conducted once half the data (N=29) is collected to assess if the planned sample size of 58 provides 95% power to detect the calculated effect size. The hypothesis will only be tested if the required sample size is confirmed to be 58 or fewer. | - We will conduct Two-way ANOVA (Face type × Condition) to test pre-to-post intervention changes in threat response. - Follow-Up Mixed-effect model analyses (Stimulus-Level):   Δendpoint = 1 + condition + order + condition*order + (1\|patient)  Where the Δendpoint is pre-to-post intervention changes in 1a) SCRs to angry male faces, 2a) SCRs to happy female faces, 1b) amygdala reactivity to angry male faces, and 2b) amygdala reactivity to happy female faces.   - We will apply paired-samples t-tests to compare a) SCRs and b) amygdala reactivity between face types at pre-training test and post-training test, respectively. | - Statistically significant results in the ANOVA interaction term will suggest the effects of DecNef conditions on physiological threat responses differ across the face types. - The statistically significant effects of condition in the mixed-effect model with the same sign with the main analyses in 1) angry male face and the inverse sign in 2) happy female faces will respectively suggest that the result in the main model is driven by changes in a) SCR and b) amygdala reactivity to that face type. - Statistically significant t-test results will suggest that threat responses are either activated or suppressed in the pre-training test or pot-training test, depending on the observed direction of changes.   See null hypothesis policy*. The SESOI will be set at 20% of the calculated effect size determined during the interim analysis. |
|  | **(SH_TPE_ 5)** Participants will show greater reduction in every PTSD symptoms cluster in the experimental condition than the control condition. | We will use the same dataset collected for testing **H_TPE_ 1**. This analysis will be conducted only when the hypothesis **H_TPE_ 1** yields statistically significant results.  Interim analyses will be conducted once half the data (N=29) is collected to calculate the SESOI. | Mixed-effect model analyses:  Δendpoint = 1 + condition + order + condition*order + (1\|patient)  Where the Δendpoint is reduction in 1) re-experiencing, 2) avoidance, and 3) hypervigilance symptoms cluster from pre-training test (-1 day) to post-training test (+ 1 week). | Positive/negative and statistically significant effects of the condition will suggest that the experimental/control condition of DecNef provides a therapeutic effect for PTSD in the corresponding symptoms cluster.  See null hypothesis policy*. The SESOI will be set at 20% of the calculated effect size determined during the interim analysis. |
|  | **(SH_TPE_ 6)** Participants with the non-dissociative subtype will show greater reduction in PTSD severity than the dissociative subtype. | Same as above. | Mixed-effect model analyses:  Δendpoint = 1 + condition + order + condition*order + subtype + (1\|patient)  Where the Δendpoint is reduction in CAPS score from pre-test (-1 day) to post-test (+ 1 week).  The subtype is coded as 1/0 for the non-dissociative/dissociative subtype. | Positive/negative and statistically significant effects of the subtype will suggest that the non-dissociative/dissociative subtype show the greater reduction in PTSD severity that the other subtype.  See null hypothesis policy*. The SESOI will be set at 20% of the calculated effect size determined during the interim analysis. |
|  | **(SH_TPE_ 7)** Across-patient variability in 1) SCRs and 2) amygdala reactivity to angry faces is correlated with across-patient variability in symptom imbalance (SI). | We will use the same dataset collected for testing **H_TPE_ 1**. Interim analyses will be conducted once half the data (N=29) is collected to assess if the planned sample size of 58 provides 95% power to detect the calculated effect size. The hypothesis will only be tested if the required sample size is confirmed to be 58 or fewer. | We will apply a mixed effect model:  Fear = 1 + SI + (SI\|patient)  Where the Fear is 1) SCRs and 2) amygdala reactivity. | Hypothesis will be supported if SI is positively and statistically significant.  See null hypothesis policy*. The SESOI will be set at 20% of the calculated effect size determined during the interim analysis. |
| **[DNF: Distress during Neurofeedback]**  Does DecNef convey weak distress/consciousness during the procedure? | **(SH_DNF_ 1)** The levels of SUDS differ significantly between the experimental and control conditions | We will use the same dataset collected for testing **H_TPE_ 1**. Interim analyses will be conducted once half the data (N=29) is collected to calculate the SESOI. | We will apply a mixed effect model:  SUDS = 1 + Condition + (1\|days) + (1\|order) + (1\|patient) | If the effects of condition are positive/negative and statistically significant, we will consider that induction of angry/happy male faces evokes more distress than those of happy/angry female faces. See null hypothesis policy*. The SESOI will be set at 20% of the calculated effect size determined during the interim analysis. |
|  | **(SH_DNF_ 2)** The dropout definition 2) is larger in the experimental condition than the control condition. | Same as above. | We will apply a Chi-square test to compare the ratio of dropout in definition 2) between experimental and control conditions. | If the dropout is significantly larger in experimental/control condition, we will consider that experimental/control session induces some distress related to the target neural representation pattern.  See null hypothesis policy*. The SESOI will be set at 20% of the calculated effect size determined during the interim analysis. |
|  | **(SH_DNF_ 3)** The ratio of participants who reported their use of conscious strategies related to 1a) visual images of angry male faces and 1b) strategies related to traumatic events in the free answer descriptions and 2) angry face in the four-forced-choice question is higher in the experimental condition than the control condition. | Same as above. | Chi-square to test if the ratios of conscious strategies differ between experimental and control conditions in the 1a) and 1b) free answer descriptions and 2) four-forced-choice question. | Significant results will suggest participants are 1) explicitly, and 2) implicitly conscious that the content for the neural representation is related to 1a) and 2) angry male faces and 1b) trauma-related stimuli.  See null hypothesis policy*. The SESOI will be set at 20% of the calculated effect size determined during the interim analysis. |
|  | **(SH_DNF_ 4)** The levels of threat responses during the successful induction periods differ between the experimental and control conditions. | Same as above. | We will use the paired-samples t-tests to compare the levels of a) SCR and b) amygdala reactivity between the experimental and control conditions. | If a) SCR and b) amygdala reactivity is higher in the experimental/control condition, we will consider that the successful induction in the experimental/control condition evokes meaningful a) peripheral and b) neural threat responses.  See null hypothesis policy*. The SESOI will be set at 20% of the calculated effect size determined during the interim analysis. |
| **[SDC: Distress during Decoder Construction]**  Does Decoder Construction convey weak distress/consciousness during the procedure? | **(SH_DDC_ 1)** Participants selectively detect angry male faces in comparison with happy female faces. | Same as above. | We will apply a Chi square test to compare the ratio of conscious trials between the trials where angry male faces were presented versus those where happy female faces were presented. | If detection rate is higher in angry male/ happy female faces, we will consider that participants selectively detect angry male/happy female faces in comparison with other faces.  See null hypothesis policy*. The SESOI will be set at 20% of the calculated effect size determined during the interim analysis. |
|  | **(SH_DDC_ 2)** The presentation of angry male faces during the decoding sessions induce larger a) SCRs or b) amygdala reactivity than the happy female faces. | Same as above. | We will apply paired t-tests between a) SCRs and b) amygdala reactivity to angry male faces and those to happy female faces during the decoding session. | If a) SCR and b) amygdala reactivity is higher to angry male/happy female faces, we will consider that the presentation of angry male/happy female faces evokes meaningful a) peripheral and b) neural threat responses.  See null hypothesis policy*. The SESOI will be set at 20% of the calculated effect size determined during the interim analysis. |

Our hypothesis posits no significant differences for **SH_BC_ 1, 2, SH_DNF_**, and **SH_DDC_**, and assumes heterogeneity in DecNef’s effects across symptom clusters for **SH_TPE_ 5**. However, to ensure comprehensive testing of all possible outcomes, we will first perform null hypothesis rejection testing to identify significant effects. Should the null hypothesis remain unrejected, we will move on to equivalence testing to evaluate whether observed effects lie within the predefined equivalence margins.

*null hypothesis policy: In case the null hypothesis cannot be rejected, we will conduct an equivalence test using an interval of [−SESOI, SESOI]. If the observed 90% confidence interval is entirely within this equivalence range, we will interpret this as evidence supporting a null effect. Conversely, if the confidence interval falls outside this range, the results will be deemed inconclusive regarding the null hypothesis.

**Baseline comparison (SH_BC_ 1**, **SH_BC_ 2)**

Hypothesis **SH_BC_ 1**: To compare the two patient groups, that is, the group completing the experimental condition first and the group completing the control condition first, on the basis of baseline characteristics, independent-sample t tests and Chi-square tests will be performed.

Hypothesis **SH_BC_ 2**: These analyses of demographics will also be used to compare completers with dropouts using definitions 1a, b), and 2 (see Evaluating consciousness, distress, and dropout with DecNef), respectively.

**Intent-to-treat analyses (SH_TPE_ 1, 2, 3a, 3b)**

The main analyses will be performed only on data from participants who complete the whole procedures. To examine whether results are biased by dropouts, supplementary intent-to-treat analyses^7^ will also be performed for the effect of DecNef. Specifically, we will apply the mixed-effects model in the main analysis (i.e., **H_TPE_ 1, 2, 3a, 3b**) to all available data, including dropouts, unless consent to use the data is withdrawn.

**Physiological threat responses to each stimulus type (SH_TPE_ 4a)**

In the main analyses (**H_TPE_ 4a**), subtraction of threat responses (SCRs and amygdala reactivity) to happy female faces from those to angry male faces will be analyzed using the mixed-effects model. The analyses **(SH_TPE_ 4a)** explore the possibility that reactivity for a specific face type, including happy female faces, may be affected by DecNef, considering that happy female faces are reinforced in the control DecNef condition. This supplementary hypothesis will also clarify whether the main effects are specifically attributable to changes in response to angry male faces in the experimental condition, as opposed to responses to happy female face or control conditions. First, a two-way ANOVA (Face type × Condition) will assess pre-to-post intervention changes in physiological threat responses. Subsequently, the same mixed-effects model from the main analyses will be applied to SCRs and amygdala reactivity for each face type independently. Finally, paired-samples t-tests will compare physiological threat responses across face types at pre-test and post-test. These steps aim to determine whether DecNef modulates physiological threat responses by normalizes hyperreactivity, rescuing suppressed responses, or influencing patterns differently across conditions.

**Analysis of DecNef effects on each PTSD symptoms cluster (SH_TPE_ 5)**

As a similar procedure succeeded in fear reduction among non-clinical samples^8,9^, the experimental condition to reinforce angry male face induction is expected to lead to main reduction of the re-experiencing symptom cluster of PTSD, which is characterized by exaggerated responses to traumatic stimuli^10^. Additionally, the control condition to reinforce happy female face induction seeks to reduce emotional numbing (part of the avoidance symptom cluster of PTSD in DSM-IV). This symptom cluster includes the inability to have positive feelings for positive events or stimuli^10^. We hypothesize that the control condition will reinstate the ability of emotionally positive stimuli to elicit positive feelings by reinforcing neural representations for a happy female face with reward. This condition is expected, in turn, to lead to main reduction of the avoidance symptom cluster of PTSD in CAPS-IV. Therefore, while we expect from our pilot study that the experimental condition is superior to the control condition in ameliorating PTSD symptoms, it is worth examining an alternative possibility that the avoidance symptom cluster will be reduced more in the control condition. To examine whether the therapeutic effect of DecNef is specific to any PTSD symptom cluster, we will perform the same mixed-effect on potential reductions in CAPS scores as would be done for general PTSD severity (**H_TPE_ 5**).

**Effect of PTSD subtype on DecNef therapeutic efficacy (SH_TPE_ 6)**

The two types of symptoms of PTSD, emotional under- and overmodulatory symptoms, have led to introduction of a novel subtype of PTSD in the DSM-5, the dissociative subtype of PTSD^10^. While the typical, non-dissociative subtype is characterized by emotional undermodulation, the dissociative subtype is characterized by emotional overmodulation^11,12^. Usually, patients with the dissociative subtype show poorer therapeutic responses in comparison with those having the non-dissociative subtype^11,12^. Therefore, in this proposed study, we will examine whether DecNef therapeutic effects are also affected by subtypes at the time of enrollment.

**Correlation between across-patient variabilities in physiological threat responses and the index of emotional modulatory state (SH_TPE_ 7)**

Recently, we proposed that PTSD symptom imbalance can explain the variability in physiological threat responses, such as amygdala reactivity, more robustly than overall PTSD severity^13^. Symptom imbalance is defined as subtraction of the avoidance symptom cluster of PTSD from the re-experiencing symptom cluster of PTSD^7^. Therefore, we will examine whether across-patient variability in symptom imbalance are accompanied by across-patient variability in “Fear” (either SCR or amygdala reactivity) using the following mixed effect model:

Fear = 1 + SI + (SI|patient)

where “SI” denotes symptom imbalance.

**Evaluating consciousness, distress, and dropout with DecNef** (Hypothesis **SH_DNF_ 1-4**)

To provide additional insights into both the conscious and physiological effects of DecNef, we will supplementary examine whether experimental and control conditions of DecNef produce differing conscious experiences.

**Hypothesis SH_DNF_ 1:** SUDS levels for each DecNef session will be analyzed using the following mixed effect model:

SUDS = 1 + Condition + (1|days) + (1|order) + (1|patient)

where “days” denotes day of the DecNef (day1, day2, or day3) and “Order” denotes the order of conditions (experimental condition first, or control condition first). If distress is systematically induced due to an underlying mechanism of DecNef, then the effect of Condition is expected to be significant.

**Hypothesis SH_DNF_ 2:** Dropout definition 2) will also be used so that we can compare the experimental condition and the control condition. We will conclude that the experimental session induces some distress related to the target neural representation pattern of trauma-related angry male faces only if the dropout rate with definition 2) is statistically higher in the experimental condition than in the control condition, which will be analyzed using a chi-square test.

**Hypothesis SH_DNF_ 3:** Conscious awareness of neural representations of trauma-related stimuli, i.e., angry male faces, during DecNef will be assessed as follows: We will first evaluate ratios of participants who reported their use of conscious strategies related to 1) visual images of angry male faces and 2) strategies related to traumatic events, including visual images of angry male faces, based on their open-ended responses, at the end of each DecNef session (see DecNef session). Categorization of strategy will be performed independently by two judges, including the doctor-in-charge. The corresponding author will categorize the strategy in case of conflict. We will use a chi-square test to examine whether ratios of conscious strategies differ between experimental and control conditions. Further, chi-square tests will be performed on the four-forced-choice question asking participants to report the content of induced neural representations during DecNef, i.e., experimental versus control. Chi-square tests will be replaced with Fisher's exact test if expected frequencies in cells are below 5.

**Hypothesis SH_DNF_ 4:** To objectively evaluate distress levels during DecNef, we will evaluate the SCR and Amygdala reactivity. We will compare levels of SCR and Amygdala reactivity between the experimental and control conditions during induction periods of DecNef trials, in which the likelihood of the target faces (i.e male for experimental and female for control) exceeds 50%. When the score is 50%, the likelihood of angry male and happy female faces are equal in that trial.

**Evaluating consciousness and distress during decoder-construction sessions** (**SH_DDC_ 1, 2**)

To provide additional insights into both the conscious and physiological experiences during decoder construction sessions, we will examine whether presentations of angry male face and happy female face during decoder construction produce different conscious and physiological experiences.

**Hypothesis SH_DDC_ 1:** To evaluate the level of conscious awareness of trauma-related angry male faces during the decoding session, we will evaluate the ratio of trials in which participants reported seeing any image beside the Mondrian patches used as masks (see Section Decoding session). Specifically, we will compare the ratio of conscious trials between trials in which angry male faces were presented versus those that presented happy female faces using a chi-square test. If the ratio is significantly higher for angry male trials than happy female trials, we will conclude that participants are more likely to become consciously aware of at least some features of trauma-related cues beyond their generic response bias to report presence of some visual images without knowing their contents. However, as we mentioned in the main manuscript (see decoding session), we will refrain from introducing more stringent tasks to evaluate specific contents of consciously perceived visual images to avoid interfering with decoder construction by inflicting bias in prior expectations.

**Hypothesis SH_DDC_ 2:** Further to evaluate the level of distress during decoder construction in terms of physiological threat responses, SCRs/amygdala reactivity for decoding sessions (11 fMRI runs) will be extracted separately for angry male faces and for happy female faces (as described in fMRI analyses/Skin conductance response analyses). Then, paired t-tests will be performed between SCRs/amygdala reactivity to the two types of stimuli to learn whether presentation of angry male faces during the decoding sessions induces meaningful physiological threat responses in participants.
